# Sex and age differences in cardiovascular risk factors and lifestyle at the onset of diabetes mellitus: a cross-sectional study in Spanish Primary Health Care

**DOI:** 10.1101/2024.09.11.24313481

**Authors:** P Vich-Pérez, B Taulero-Escalera, V García-Espinosa, L Villanova-Cuadra, P Regueiro-Toribio, I Sevilla-Machuca, J Timoner-Aguilera, M Martínez-Grandmontagne, T Abós-Pueyo, C Álvarez-Hernandez-Cañizares, G Reviriego-Jaén, A Serrano-López-Hazas, I Gala-Molina, M Sanz-Pascual, MA Salinero-Fort the LADA-PC consortium

## Abstract

**Aims:** To describe the main characteristics of patients recently diagnosed with diabetes mellitus in terms of comorbidities, cardiovascular risk factors and healthy lifestyle by sex and age group.

**Methods:** A cross-sectional, multicenter, observational study of 681 patients aged >30 years diagnosed with diabetes mellitus in the previous 4 years was performed. The patients were treated in primary care centers in Madrid (Spain). The variables were obtained from their electronic medical records, physical examination, complete analysis, and lifestyle questionnaires.

**Results:** The main comorbidities were: hypercholesterolemia (64.4%; 95% CI, 60.6-68.2), hypertension (55.2%; 95% CI, 51.3-59.1), obesity (58.9%; 95% CI, 55.2-62.6), metabolic syndrome (58.5%; 95% CI, 54.6-62.5); and hypertriglyceridemia (25.3%; 95% CI, 21.9-28.7). Despite being newly diagnosed, 7.6% (95% CI, 5.4-9.8) had microalbuminuria, and 10.3% (95% CI, 8.0-12.6) cardiovascular disease. The main unhealthy lifestyles were: low physical activity (52%; 95% CI, 48.1-55.9), alcohol consumption (47.7%; 95% CI, 44.0-51.5) and smoking (19.2%; 95% CI, 16.2-22.3). Compared with men, women had more morbid obesity (9.7% vs 4.6%, p=.014), worse lipid profile (total cholesterol: 184 (IQR, 158-207) vs. 165 (IQR, 144-192), p<.01), less treatment with metformin (74.8% vs. 84.4%, p<.01) and antiplatelet agents (8.1% vs.18.6%, p<.01), but women had fewer comorbidities. Patients with a high educational level (OR= 1.90, 95% CI, 1.28-2.81)) and those >60 years (OR= 1.49; 95% CI, 1.01-2.21) were more adherent to the Mediterranean diet, and the older ones did less intense exercise (OR= 0.34, 95% CI, 0.16-0.75). Normal blood pressure was associated with Mediterranean diet (OR= 1.52; 95% CI, 1.05-2.21) and high physical activity (OR= 4.03; 95% CI, 1.69-9.61); and body mass index was inversely associated with physical activity (OR= 0.92; 95% CI, 0.85-0.99).

**Conclusions:** Patients newly diagnosed with diabetes mellitus have crucial cardiovascular risk factors and comorbidities at the onset of the disease. These can be modified through a healthy lifestyle.

## Introduction

Diabetes mellitus (DM) is one of the most common metabolic disorders worldwide [1], with increasing prevalence in most countries. Type 2 DM (T2DM) affects approximately 90% of people with DM worldwide. Within Europe, Spain has one of the highest prevalence values. The 2012 study found a prevalence of T2DM in Spain of 13.8%, with almost half of them (6%) unaware that they had the disease [2].

In 1980, the World Health Organization (WHO) published the first recommendations for the diagnosis and classification of DM, which were updated in 1985 [3]. The diagnostic criterion for DM was established as a fasting plasma glucose level ≥140 mg/dl. Beginning in 1998, the WHO established a fasting plasma glucose level ≥126 mg/dl as the new diagnostic criterion for diabetes [4]. The application of these new diagnostic criteria should reduce the prevalence of complications associated with DM at the time of diagnosis. However, information on the prevalence of chronic complications (e.g. cardiovascular events, cerebrovascular events, nephropathy and neuropathy) in newly diagnosed DM remains scarce.

One of the main reasons for the high prevalence of T2DM today is the increased frequency of unhealthy lifestyles. In fact, T2DM can be largely prevented by adopting a healthy lifestyles [5,6]; it should also be diagnosed early and treated using a multifactorial approach. This has a direct impact on preventing complications, reducing mortality, and increasing healthcare costs. A healthy lifestyle encompasses different aspects, such as diet, regular physical exercise, enjoyment of leisure time, good interpersonal relationships, self-esteem, proper rest, stress management, and avoidance of toxic substances and pollutants. According to ADA recommendation, diet is an essential component of the prevention and treatment of T2DM [7]. There is abundant evidence that adhering to a Mediterranean diet is one of the most likely ways to prevent cardiovascular events in people with diabetes [8]. The PREDIMED study provided scientific data showing that the Mediterranean diet is a useful pattern for the primary prevention of cardiovascular disease (CVD) in people considered to be at high risk. It has proven effective at preventing myocardial infarction, cerebrovascular accident, and cardiovascular mortality. In addition, people following this diet had a lower incidence of other diseases, such as T2DM, metabolic syndrome, peripheral arterial disease, atrial fibrillation, arterial hypertension, cognitive impairment, and breast cancer [9,10]. Furthermore, among participants with T2DM, a Mediterranean diet supplemented with extra virgin olive oil can delay the prescription of new hypoglycemic medications [11].

Regarding physical activity in people with T2DM, the American Diabetes Association (ADA) recommends at least 150 minutes per week of moderate-intense aerobic physical exercise spread over at least 3 sessions and avoiding two consecutive days without exercise [12,13]. One of the most recent Spanish studies on healthy lifestyles in T2DM patients, showed that less than 23% of patients adhered closely to the Mediterranean diet, and that only 10% adhered to all healthy lifestyle habits (balanced diet, regular physical activity, not smoking, adequate weight control, and emotional well-being) [14].

In other countries, adherence to healthy lifestyles in T2DM patients is variable, although adherence to each additional healthy lifestyle factor was associated with a reduced relative risk of 32% (95% CI, 28-36) for T2DM and 21% (95% CI, 15-26) for mortality [15].

To our knowledge, few lifestyle studies address the early stages of DM in adults, when it is essential to implement an appropriate lifestyle.

The aim of this study was to describe the main characteristics of patients with newly diagnosed DM regarding cardiovascular risk factors, diabetes-related complications, and healthy lifestyle and to compare possible differences according to sex and age group.

## Materials and Methods

This work is part of a larger study (LADA-PC) to develop and validate a clinical score to identify patients at high risk of latent autoimmune diabetes in adults (LADA) among people with newly diagnosed DM. The characteristics of this work have been previously published [16] and recruitment is ongoing until the predefined sample size is reached.

Briefly, this cross-sectional, multicenter, descriptive, and observational study was carried out in 681 patients recently diagnosed with DM in primary health care centers of the Madrid Health Service SERMAS (Spain).

The inclusion criteria were as follows: diagnosis of DM after 30 years of age recorded in the electronic medical record (EMR), and diagnosis of DM within the last four years with a duration of illness of at least six months.

The exclusion criteria were as follows: gestational diabetes, homebound status, and physical or psychological inability to complete the interview or undergo a brief examination and analysis.

A total of 270 professionals (general practitioners and nurses) from 12 health centers were given a list of patients who met the inclusion criteria. The professionals were instructed to consecutively include the maximum number of patients according to avoid selection bias (convenience recruitment). They conducted interviews and collected anthropometric, clinical, analytical, and psychosocial data. These data were obtained from the blood analysis during the previous 6 months, data from the EMR and physical examination findings.

The study was approved by the Research and Medicines Ethics Committee and the Central Research Commission of the Primary Care Management of Madrid with reference number “CODE: JCAH/PVP/LVP/LVP/LVP/LVP/LADA/2019/1. VERSION: March 4, 2019”. All participants in the present study were enrolled from 03/30/2022 to 08/30/2023.

All data were treated confidentially and anonymized. Personal identificative data were dissociated in the electronic data collection notebook, therefore, it was not possible for people outside the project to identify the patients (compliance with Organic Law 3/2018, of December 5, on the Protection of Personal Data and Guarantee of Digital Rights). Below we address the timing of each phase. At the inclusion visit, participants were confirmed as meeting the diagnostic criteria for DM of the American Diabetes Association (ADA). After signing the informed consent form, patients were called for a complete blood test if they had not undergone one during the previous six months (complete blood count, complete biochemical analysis, HbA1c and microalbuminuria). At a subsequent visit, 7 to 30 days after baseline, all study variables were completed, and patients were informed of the results of the analysis.

### Variables

ADA diagnostic criteria for DM: 1. Cardinal symptoms + Fasting Plasma Glucose (FPG) ≥ 200 mg/dl. 2. HbA1c ≥6.5% on at least two occasions. 3. Two FPGs levels ≥126 mg/dl. 4. FPG 2 hours after the oral glucose tolerance test (2-h OGTT) with 75 g glucose ≥200 mg/dl on two occasions. 5. Two abnormal results from the same sample, Hb A1c ≥ 6.5% and FPG ≥126 mg/dl at least once each [17].

Some of the variables required were completed from the EMRs, such as previous chronic pathologies, therapies, clinical history, essential blood and biochemical analyses, and anthropometric data from the previous 6 months. An in-person visit was required to collect other variables, such as complete biochemical analysis, height, weight, body mass index (BMI [weight (kg) / height^2^ (m)]), waist circumference and family clinical history. Lifestyle and sociodemographic questionnaires were also administered.

### Variable definitions

#### Metabolic syndrome

Metabolic syndrome was defined according to the National Cholesterol Education Program Adult Treatment Panel III as the presence of at least three of the following components: 1) abdominal circumference ≥102 cm in men and ≥88 cm in women; 2) triglycerides ≥150 mg/dl; 3) blood pressure ≥130/85 mm/Hg or established arterial hypertension; 4) HDL cholesterol <40 mg/dl in men and <50 mg/dl in women; 5) fasting plasma glucose (FPG) 110 to 126 mg/dl (6.11 to 6.99 mmol/L) or a diagnosis of DM [18].

#### Cardiovascular disease (CVD)

CVD was defined as cerebrovascular disease, coronary heart disease or peripheral arterial disease.

#### Smoking habit

We considered three levels of smoking habit: active smoker (current smokers, either daily or occasionally), never smoker, and ex-smoker (not smoked for at least 1 year).

#### Alcohol consumption

Weekly units of alcohol were recorded as the sum of the units consumed. One unit (10 g of alcohol) is equivalent to one glass of wine (100 cc), one of beer (200 cc), half a glass of vermouth (50 cc) or half a measure of whisky or cognac (25 cc). The volume was measured as follows: small glass, 125 ml; medium glass, 200 ml; large glass, 250 ml; glass of mixed drink, 50 ml; bottle of beer, 200 ml; bottle of wine, 750 ml. The electronic booklet tool included a calculator that calculates the daily and weekly alcohol consumption and categorized patients as heavy drinkers (>40 g of alcohol/day for men, >24 g for women), drinkers but not at risk (>0 and <40 g of alcohol/day for men, >0 and <24g for women), and nondrinkers.

#### Educational level

We considered the following levels: no or incomplete primary education, primary education, junior and senior high school, basic and advanced vocational training and university studies.

#### Physical activity level

A reduced scale of the International Physical Activity Questionnaire (IPAQ) was used [19] with the following categories: low (not performing any physical activity or the activity performed is not sufficient to meet IPAQ category 2 or 3); moderate (≥3 sessions/week of vigorous physical activity for at least 20 minutes per day or ≥5 sessions/week of moderate physical activity and/or walking for at least 30 minutes per day; or ≥5 sessions/week of any combination of walking and/or moderate and/or vigorous physical activity); and high (≥3 sessions/week of vigorous physical activity for at least 60 minutes per day or ≥7 sessions/week of any combination of walking and/or moderate and/or vigorous-intensity physical activity).

#### Mediterranean diet

A modified version of the Mediterranean Diet Adherence Screener (MEDAS) questionnaire was used to analyze adherence to the Mediterranean diet (a score of 11-14 points indicated high adherence). The modification concerns wine consumption which has been reduced to two glasses per week instead of seven as in the original MEDAS. Given that the variable alcohol consumption can be classified by risk based on the type of drink and grams/week, not only considering the number of glasses, it was easily recorded.

This questionnaire has been found to be a valid tool for rapid assessment of adherence and may be useful in clinical practice [20].

#### Polypharmacy

Polypharmacy was defined as the regular intake of five or more medications per day, as reported in most studies [21].

### Statistical analysis

Descriptive statistics of the demographic and clinical characteristics of the patients included in the study were performed and stratified by sex and age. Continuous quantitative variables were summarized as the mean and standard error (LDL cholesterol and microalbuminuria) or median and interquartile range when necessary. Qualitative variables were summarized as absolute frequencies or relative frequencies (%). The Student t test or the Mann Whitney U test for nonnormal distributions was used to compare subgroups of quantitative variables. The chi-squared test or z test for the comparison of proportions was used to compare qualitative variables.

Two logistic regression models were developed to investigate independent associations of age, sex, and educational level with high adherence to the Mediterranean diet (MEDAS-14 score greater than 10) and with high physical activity, respectively. The adjusting covariates (smoking, alcohol consumption, hypertension, cardiovascular disease [coronary heart disease, cerebrovascular disease, peripheral arterial disease], heart failure, atrial fibrillation, BMI, metformin, sulfonylureas, insulin, SGLT-2 inhibitors, DPP-4 inhibitors, GLP-1 RA, anticoagulants, and beta-blockers were included in the maximum model. The main variables were entered using the enter method, and the covariates were entered using the backward stepwise method (LR). The variation of the OR of the main variables with and without the suspected confounding variable was tested to avoid confounding. Collinearity between variables was also analyzed.

All the statistical analyses were performed with SPSS for Windows version 26.0 (IBM SPSS, IBM Corp, Armonk, NY, USA: IBM Corp.). A 2-tailed p value <.05 was considered statistically significant.

## Results

### Baseline characteristics

Of the newly diagnosed patients, 741 were contacted and a high participation rate of 93.5% was achieved, with 693 agreeing to participate in the study. Unfortunately, 12 participants had to be excluded due to missing key analysis variables, resulting in a final study sample of 681 patients. The median age was 63 years (IQR, 56-71) and the majority were men (56.1%, n= 382). The median age at diagnosis was 59 years (IQR, 53-67).

The main diagnostic method was FPG ≥126 mg/dl on two occasions (42.6%; 95% CI, 38.9-46.3).

Most participants had a basic educational level (21.4% [95% CI, 18.2-24.4] primary education and 19.4% [95% CI, 16.4-22.4] junior high school education).

The predominant comorbidities were hypercholesterolemia (64.4%; 95% CI, 60.6-68.2), obesity (58.9%; 95% CI, 55.2-62.6), metabolic syndrome (58.5%; 95% CI, 54.6-62.5) and hypertension (55.2%; 95% CI, 51.3-59.1). Followed by a quarter who had hypertriglyceridemia (25.3%; 95% CI, 21.9-28.7). Although the patients were newly diagnosed, they already had conditions associated with diabetes mellitus, such as microalbuminuria (7.6%; 95% CI, 5.4-9.8), chronic kidney disease (CKD) (3.7%; 95% CI, 2.2-5.2), retinopathy (1.6%; 95% CI, 0.1-2.6), and neuropathy (1.5%; 95% CI, 0.1-2.5).

These patients had high biochemical values at baseline (Table 1). The improvement in median of HbA1c from the date of diagnosis to the baseline visit was clinically relevant (-0.6 percentage units).

**Table 1.**
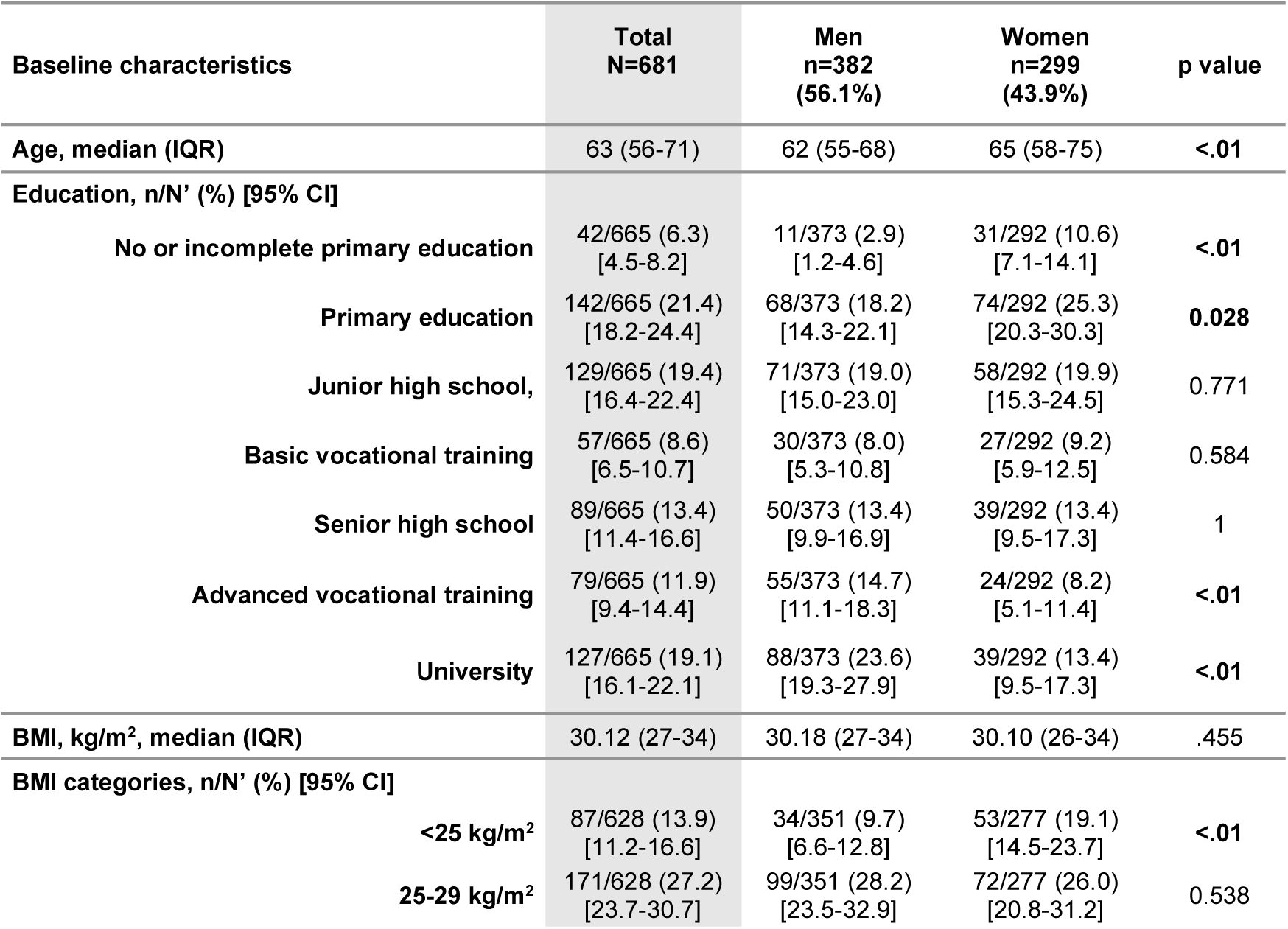
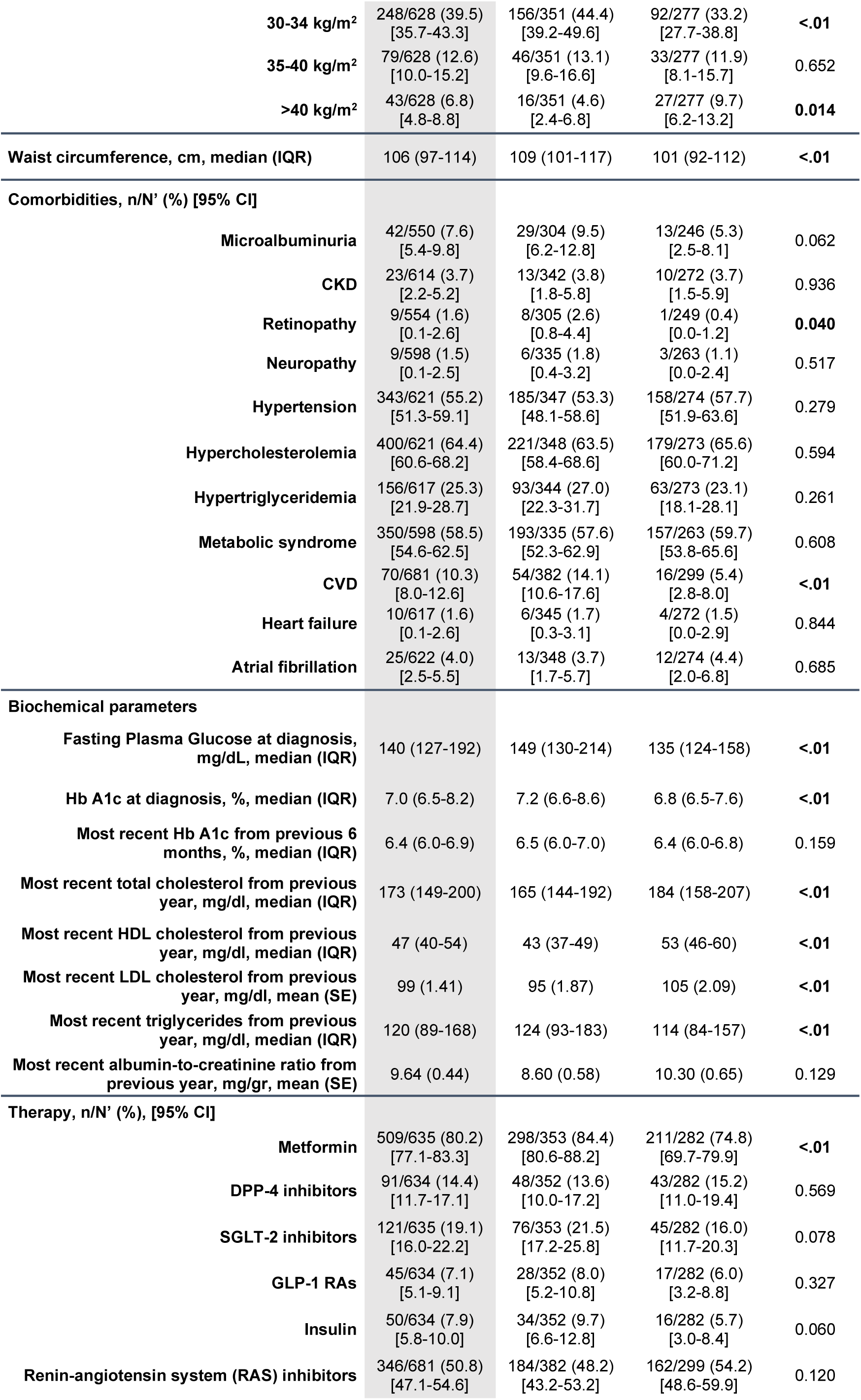
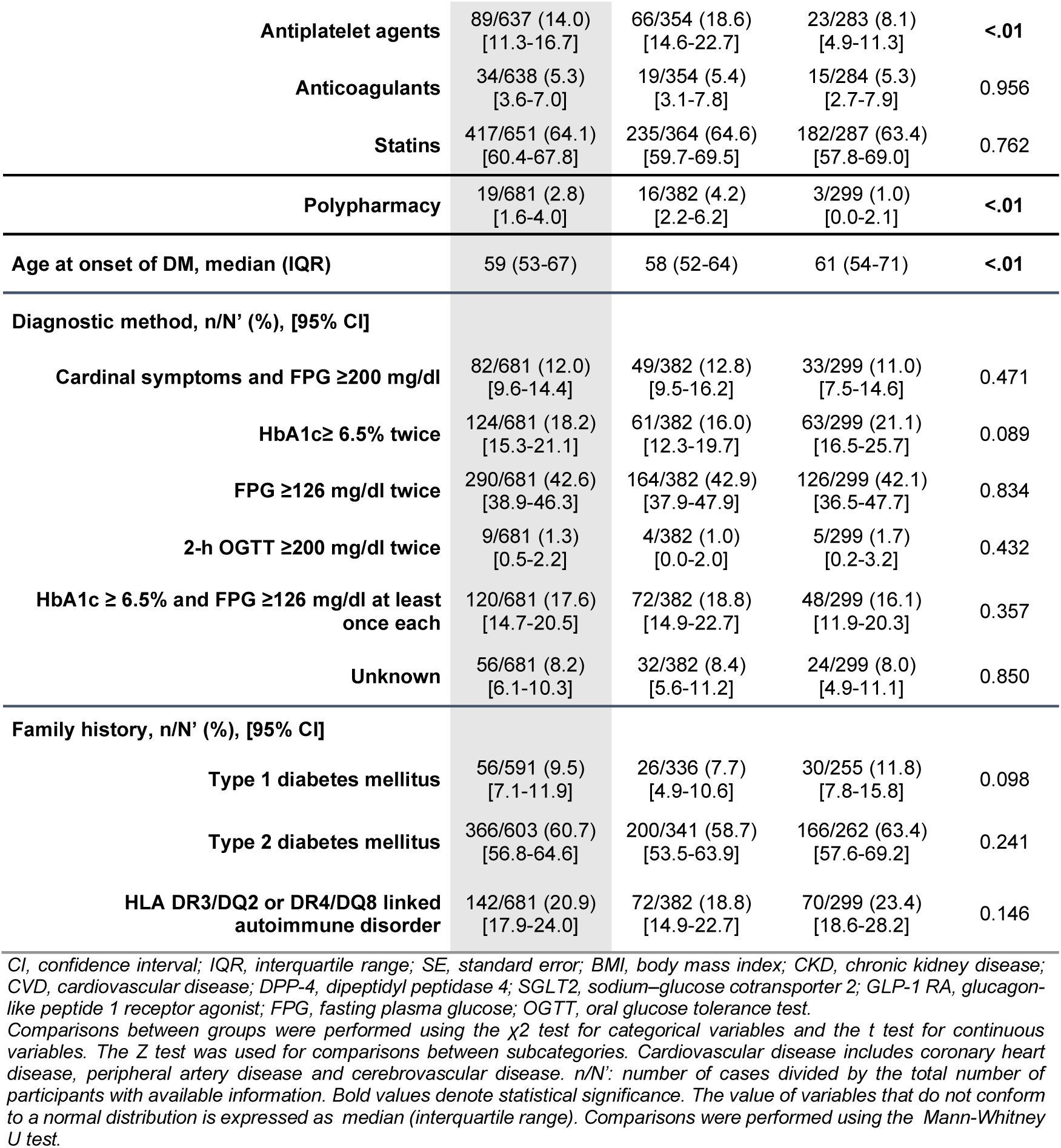
Baseline characteristics of patients with newly diagnosed DM stratified by sex.

The most frequently used drugs were metformin (80.2%; 95% CI, 77.1-83.3), statins (64.1%; 95% CI, 60.4-67.8) and renin-angiotensin system (RAS) inhibitors (50.8%; 95% CI, 47.1-54.6). Among the new classes of antidiabetic drugs, SGLT2 inhibitors were the treatment of choice (19.1%; 95% CI, 16.0-22.2) (Table 1).

### Baseline characteristics: analysis according to sex

Women were significantly older than men (65 [IQR, 58-75] vs. 62 [IQR, 55-68] years, p<.01), with a higher age at onset of DM (61 [IQR, 54-71] vs. 58 [IQR, 52-64] years, p<.01). Women more frequently had lower educational levels: no or incomplete primary education (10.6% women vs. 2.9% men, p<.01) and primary education (25.3% women vs. 18.2% men, p=0.028). Both groups had a similar median BMI. However, the distribution of BMI categories differed between the sexes. Women were more prevalent in the <25 kg/m^2^ category (19.1% vs. 9.7%, p<.01) and >40 kg/m^2^ category (9.7% vs. 4.6%, p=0.014). However, results were higher for men in the 30-34 kg/m^2^ range (44.4% vs. 33.2%, p<.01).

Comorbidities were more common in men, with a predominance of classic complications of diabetes mellitus such as retinopathy (2.6% vs. 0.4%, p=0.040) and CVD (14.1% vs. 5.4%, p<.01), which were already present at onset of DM. In the biochemical analysis, higher lipid profile values were recorded for women, as follows; total cholesterol (184 [IQR, 158-207] vs. 165 [IQR, 144-192], p<.01), LDL cholesterol (105 [SE, 2.09] vs. 95 [SE, 1.87], p<.01) and HDL cholesterol (53 [IQR, 46-60] vs. 43 [IQR, 37-49], p<.01). Triglycerides level, in contrast, were higher in men (124 [IQR, 93-183] vs. 114 [IQR, 84-157], p<.01). Both, FPG levels and HbA1c values at diagnosis were lower in women than in men (6.8 [IQR, 6.5-7.6] vs. 7.2 [IQR, 6.6-8.6], p<.01) for HbA1c and (135 [IQR, 124-158] vs. 149 [IQR, 130-214], p<.01) for FPG levels). In terms of therapy, men were more likely than women to be treated with metformin (84.4% vs. 74.8%, p<.01) and antiplatelet agents (18.6% vs. 8.1%, p<.01). In addition, men were significantly more polymedicated than women (4.2% men vs 1% women, p <.01).

### Lifestyle characteristics

At baseline, analysis of the lifestyle of patients with newly diagnosed DM (Table 2) showed that most had low levels of physical activity (52%; 95% CI, 48.1-55.9). Regarding smoking habits, 41.7% (95% CI, 48.4-56.2) were nonsmokers, 39.1% (95% CI, 35.3-42.9) were ex-smokers, and 19.2% (95% CI, 16.2-22.3) were active smokers. Regarding alcohol consumption, 52.3% (95% CI, 48.4-56.2) of the participants were teetotal, 42.9% (95% CI, 39.0-46.8) were nonrisk drinkers, and 4.8% (95% CI, 3.1-6.5) were heavy drinkers.

**Table 2.** Lifestyle characteristics at baseline in patients with newly diagnosed DM, overall and stratified by sex.

| Lifestyle characteristics | Total<br>N=681 | Men<br>n=382<br>(56.1%) | Women<br>n=299<br>(43.9%) | p value |
| --- | --- | --- | --- | --- |
| **Physical activity, n/N' (%) [95% CI]** |  |  |  |  |
| Low | 332/639 (52.0)<br>[48.1-55.9] | 175/356 (49.2)<br>[44.0-54.4] | 157/283 (55.5)<br>[49.7-61.3] | 0.113 |
| Moderate | 276/639 (43.2)<br>[39.2-46.8] | 160/356 (44.9)<br>[39.7-50.1] | 116/283 (41.0)<br>[35.3-46.7] | 0.323 |
| High | 31/639 (4.9)<br>[3.2-6.6] | 21/356 (5.9)<br>[3.4-8.3] | 10/283 (3.5)<br>[1.4-5.6] | 0.154 |
| **Smoking, n/N' (%) [95% CI]** |  |  |  |  |
| Active smoker | 123/640 (19.2)<br>[16.2-22.3] | 87/356 (24.4)<br>[19.9-28.9] | 36/284 (12.7)<br>[8.8-16.6] | <.01 |
| Ex smoker | 250/640 (39.1)<br>[35.3-42.9] | 161/356 (45.2)<br>[40.0-50.4] | 89/284 (31.3)<br>[25.9-36.7] | <.01 |
| Nonsmoker | 267/640 (41.7)<br>[48.4-56.2] | 108/356 (30.3)<br>[34.6-44.8] | 159/284 (56.0)<br>[62.6-73.6] | <.01 |
| **Alcohol, n/N' (%) [95% CI]** |  |  |  |  |
| Heavy drinker | 30/629 (4.8)<br>[3.1-6.5] | 27/350 (7.7)<br>[4.9-10.5] | 3/279 (1.1)<br>[0.1-2.3] | 0.040 |
| Drinker, but not risky | 270/629 (42.9)<br>[39.0-46.8] | 184/350 (52.6)<br>[47.4-57.8] | 86/279 (30.8)<br>[25.4-36.2] | <.01 |
| Teetotal | 329/629 (52.3)<br>[48.4-56.2] | 139/350 (39.7)<br>[34.6-44.8] | 190/279 (68.1)<br>[62.6-73.6] | 0.016 |
| **Alcohol units per week, median (IQR)\*** | 0.0 (0-4) | 1.0 (0-8) | 0.0 (0-0.8) | <.01 |
| **MEDAS Score<sup>o</sup>, n/N' (%) [95% CI]** |  |  |  |  |
| Low adherence, 0-5 | 39/631 (6.2)<br>[0.4-8.1] | 31/352 (8.8)<br>[5.8-11.8] | 8/279 (2.9)<br>[0.0-4.9] | <.01 |
| Medium adherence, 6-10 | 440/631 (69.7)<br>[66.1-73.3] | 240/352 (68.2)<br>[63.3-73.1] | 200/279 (71.7)<br>[66.4-77.0] | 0.341 |
| High adherence, ≥11 | 152/631 (24.1)<br>[20.8-27.4] | 81/352 (21.2)<br>[16.9-25.5] | 71/279 (25.4)<br>[20.3-30.5] | 0.215 |
| **14-MEDAS Questionnaire, n/N' (%) [95% CI]** |  |  |  |  |
| Olive oil as main source of fat | 607/637 (95.3)<br>[93.7-96.9] | 337/355 (94.9)<br>[92.6-97.2] | 270/282 (95.7)<br>[93.3-98.1] | 0.636 |
| Consumption of more than 3 tablespoons of olive oil per day | 312/636 (49.1)<br>[45.2-53.0] | 174/355 (45.5)<br>[40.3-50.7] | 138/281 (49.1)<br>[43.3-55.0] | 0.367 |
| Consumption of vegetables per day, more than 1 portion | 347/636 (54.6)<br>[50.7-58.5] | 172/355 (48.5)<br>[43.3-53.7] | 175/281 (62.3)<br>[56.6-68.0] | <.01 |
| Consumption of fruits per day, more than 2 | 392/635 (61.7)<br>[57.9-65.5] | 191/354 (54.0)<br>[48.8-59.2] | 201/281 (71.5)<br>[66.2-76.8] | 0.015 |
| Consumption of red meat per day, less than 1 | 460/634 (72.6)<br>[69.1-76.1] | 249/354 (70.3)<br>[65.5-75.1] | 211/280 (75.4)<br>[70.4-80.4] | 0.152 |
| Consumption of butter per day, less than 1 | 465/635 (73.2)<br>[69.8-76.7] | 254/354 (71.8)<br>[67.1-76.5] | 211/281 (75.1)<br>[70.0-80.2] | 0.350 |
| Consumption of sugary drinks per day, less than 1 | 437/636 (68.7)<br>[65.1-72.3] | 237/355 (66.8)<br>[61.9-71.7] | 200/281 (71.2)<br>[65.9-76.5] | 0.233 |
| Consumption of wine per week, more than 2 glasses | 98/635 (15.4)<br>[12.6-18.2] | 83/354 (23.4)<br>[19.0-27.8] | 15/281 (5.3)<br>[2.7-7.9] | 0.032 |
| Consumption of legumes per week, more than 2 portions | 369/635 (58.1)<br>[54.3-61.9] | 214/354 (60.5)<br>[55.4-65.6] | 155/281 (55.2)<br>[49.4-61.0] | 0.179 |
| Consumption of fish per week, more than 2 | 381/634 (60.1)<br>[56.3-63.9] | 207/354 (58.5)<br>[53.4-63.6] | 174/280 (62.1)<br>[56.4-67.8] | 0.358 |
| **Consumption of sweets per week, less than 3** | 449/634 (70.8)<br>[67.3-74.3] | 239/353 (67.7)<br>[62.8-72.6] | 210/281 (74.7)<br>[69.6-79.8] | **0.053** |
| **Consumption of nuts per week, at least 1** | 347/635 (54.6)<br>[50.7-58.5] | 207/354 (58.5)<br>[53.4-63.6] | 140/281 (49.8)<br>[44.0-55.7] | **0.029** |
| **Consumption preferably of white meat per week** | 516/635 (81.3)<br>[78.3-84.3] | 265/354 (74.9)<br>[70.4-79.4] | 251/281 (89.3)<br>[85-7-92.9] | **<.01** |
| **Consumption of sautéed vegetables per week, more than once** | 403/635 (63.5)<br>[59.8-67.2] | 224/354 (63.3)<br>[58.3-68.3] | 179/281 (63.7)<br>[58.1-69.3] | 0.917 |

CI, confidence interval; IQR, interquartile range; MEDAS, Mediterranean diet adherence screener; Comparisons between groups were performed using the $\chi^2$ test for categorical variables and the t test for continuous variables. The Z test was used for comparisons between subcategories. Diet is considered Mediterranean if the score on the 14-MEDAS is between 11-14 points. Heavy drinker: consumption >40 g alcohol/day in men, >24 g alcohol/day in women. n/N: number of cases divided by total number of participants with available information. Bold values denote statistical significance. The value of variables that do not conform to a normal distribution is expressed as in median (interquartile range). Comparisons were performed using the Mann-Whitney U test. ♦ MEDAS Score categories: Low adherence=0-5 points; Medium adherence=6-10; High adherence=11-15.

Adherence to the Mediterranean diet was moderate in 69.7% (95% CI, 66.1-73.3) of patients, high in 24.1% (95% CI, 20.8-27.4), and low in 6.2% (95% CI, 0.4-8.1). Most patients with newly diagnosed DM consumed olive oil as their main source of fat (95.3%; 95% CI, 93.7-96.9), and 49.1% (95% CI, 45.2-53.0) consumed at least the four daily recommended tablespoons of olive oil. Regarding meat consumption, 72.6% (95% CI, 69.1-76.1) ate less than one portion of red meat per day, and 81.3% (95% CI, 78.3-84.3) preferred white meat.

### Lifestyle: differences by sex

In the analysis by sex distribution (Table 2), men were predominant among active smokers (24.4% vs. 12.7%, p<.01) and among ex-smokers (45.2% vs. 31.3%, p<.01). There was a predominance of men over women in alcohol consumption for heavy drinker (7.7% vs. 1.1%, p=0.040), and drinker, but not risky (52.6% vs. 30.8%, p<0.01).The median [IQR] number of alcohol units per week was higher in men than in women (1 [0–8] vs. 0 [0-0.8], p<.01).

Men were less adherent to the Mediterranean diet (8.8% of low MEDAS score vs. 2.9%, p<.01), consuming significantly less fruit and vegetables per day than women. They also consumed less white meat (89.3% of women vs. 74.9% of men, p<.01).

Regarding snack consumption, women consumed fewer commercial sweets per week than men did. In addition, men consumed more nuts per week than women (58.5% vs. 49.8%, p=0.029).

### Analysis according to sex and age groups

Table 3 shows the differences in age groups between the sexes. Men aged >60 years had a significantly higher prevalence of CKD than those aged ≤60 years (1.3% ≤60 vs. 5.9% >60, p=0.028), in contrast to women. However, men ≤60 years had significantly higher median [IQR] values than men >60 years, in contrast to women, for each of the following variables: BMI (31.1 [27.8-34.3] kg/m^2^ ≤60 vs. 29.8 [26.9-32.9] kg/m^2^ >60, p=.002), FPG (169 [133–250] mg/dl ≤60 vs. 138 [127–177] mg/dl >60, p<.01) and HbA1c in previous 6 months (7.8% [6.8-10.3] ≤60 vs. 7.0% [6.5-7.9] >60, p<.01), total cholesterol (169 [153–195] mg/dl ≤60 vs. 160 [138–187] >60, p=0.022) and triglycerides (146 [100–212] mg/dl ≤60 vs. 115 [87–148] mg/dl >60, p<.01), and higher frequency of treatment with GLP-1 RA (12.6% ≤60 vs. 2.9% >60, p<.01) and treatment with insulin (12.6% ≤60 vs. 5.8% >60, p=0.022), cardinal symptoms and FPG >200 mg/dl as diagnostic criteria (16.7% ≤60 vs. 9.6% >60, p= 0.041), family history of T2DM (67.5% ≤60 vs. 51.1% >60, p=0.002), and family history of autoimmune disorders (24.1% ≤60 vs. 14.4% >60, p=0.016). On the other hand, older women were significantly more likely to use renin-angiotensin system inhibitors therapy than younger women (66.7% vs. 39.2%, p<.01), in contrast to men.

**Table 3.**
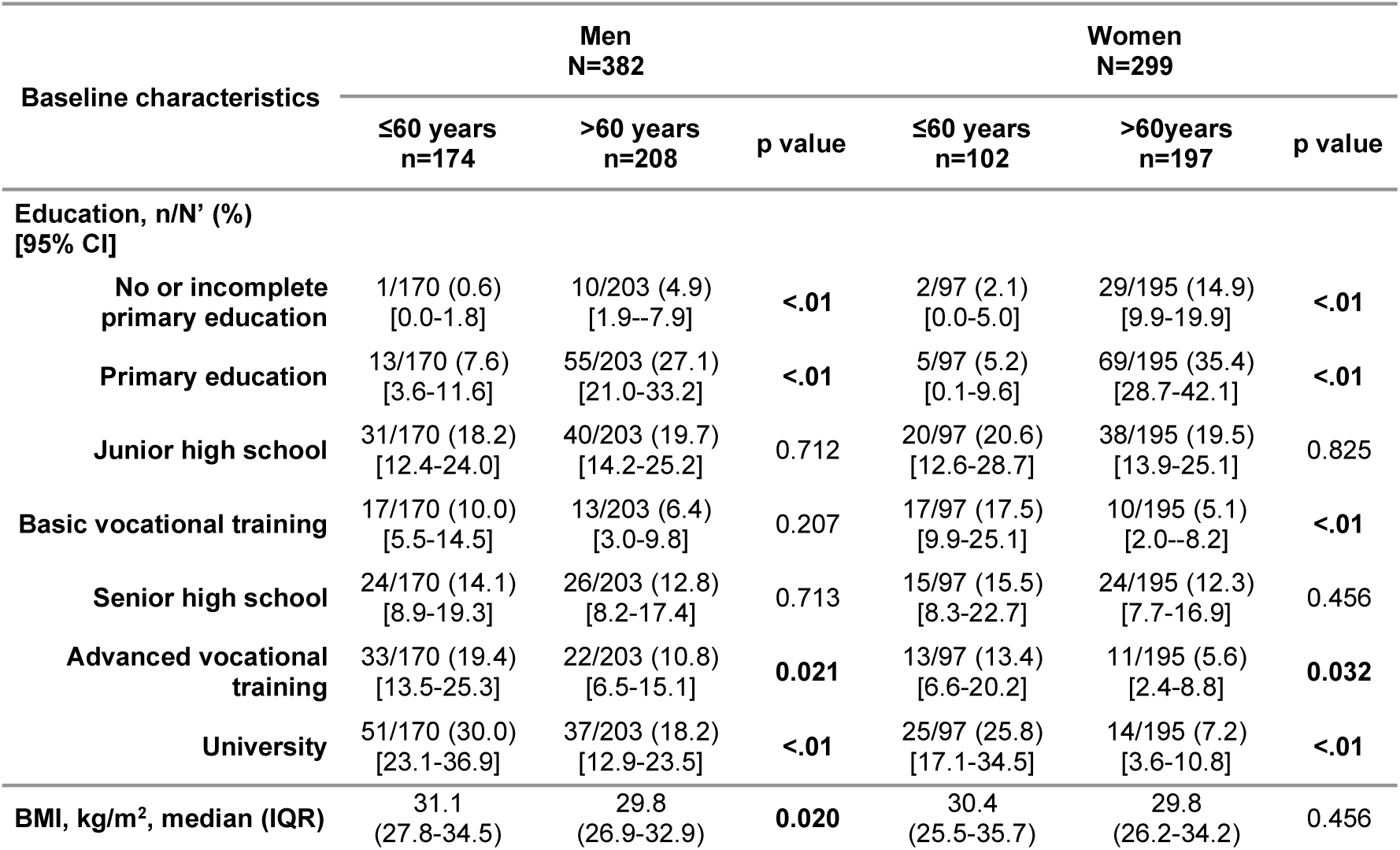
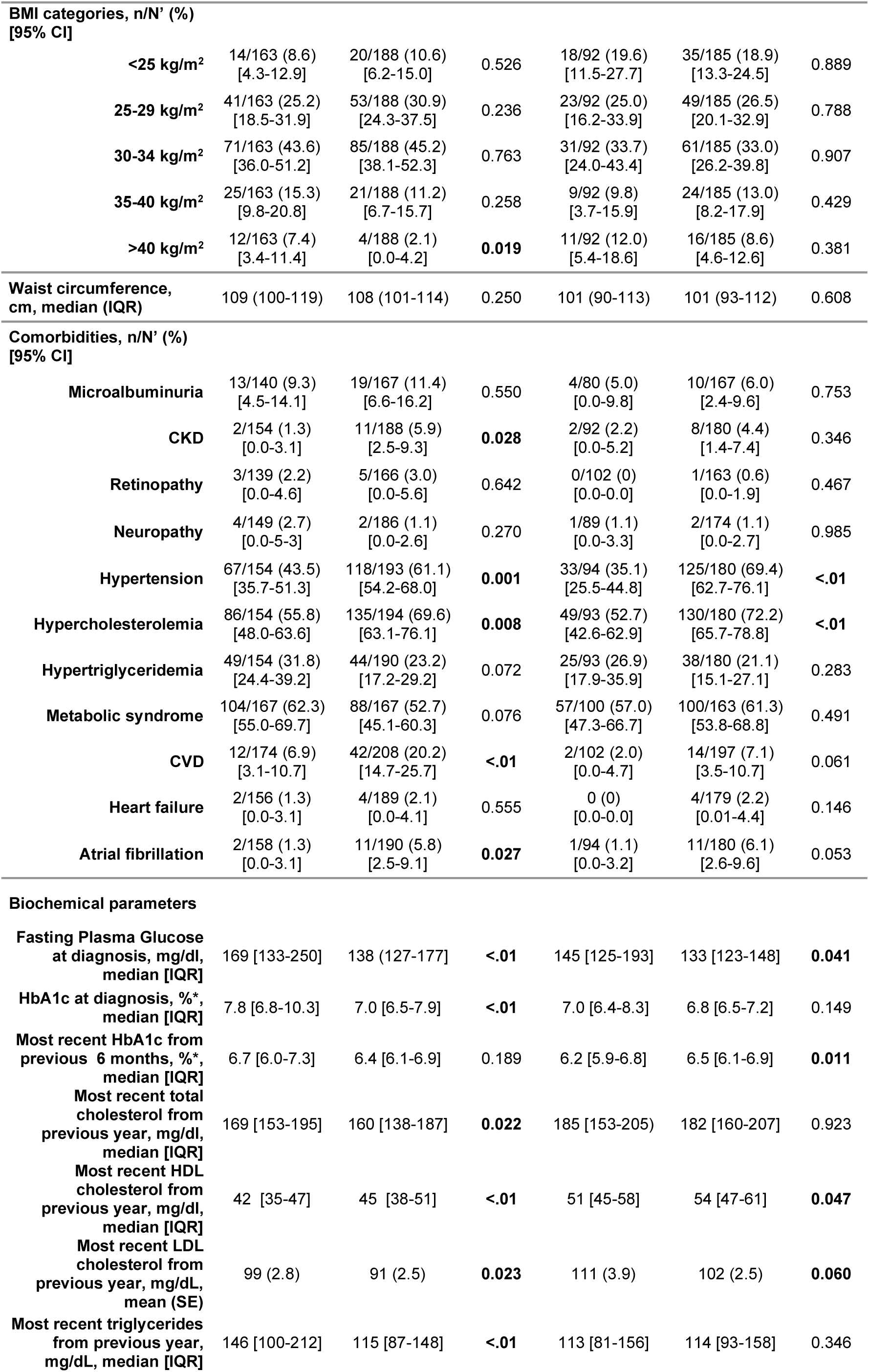
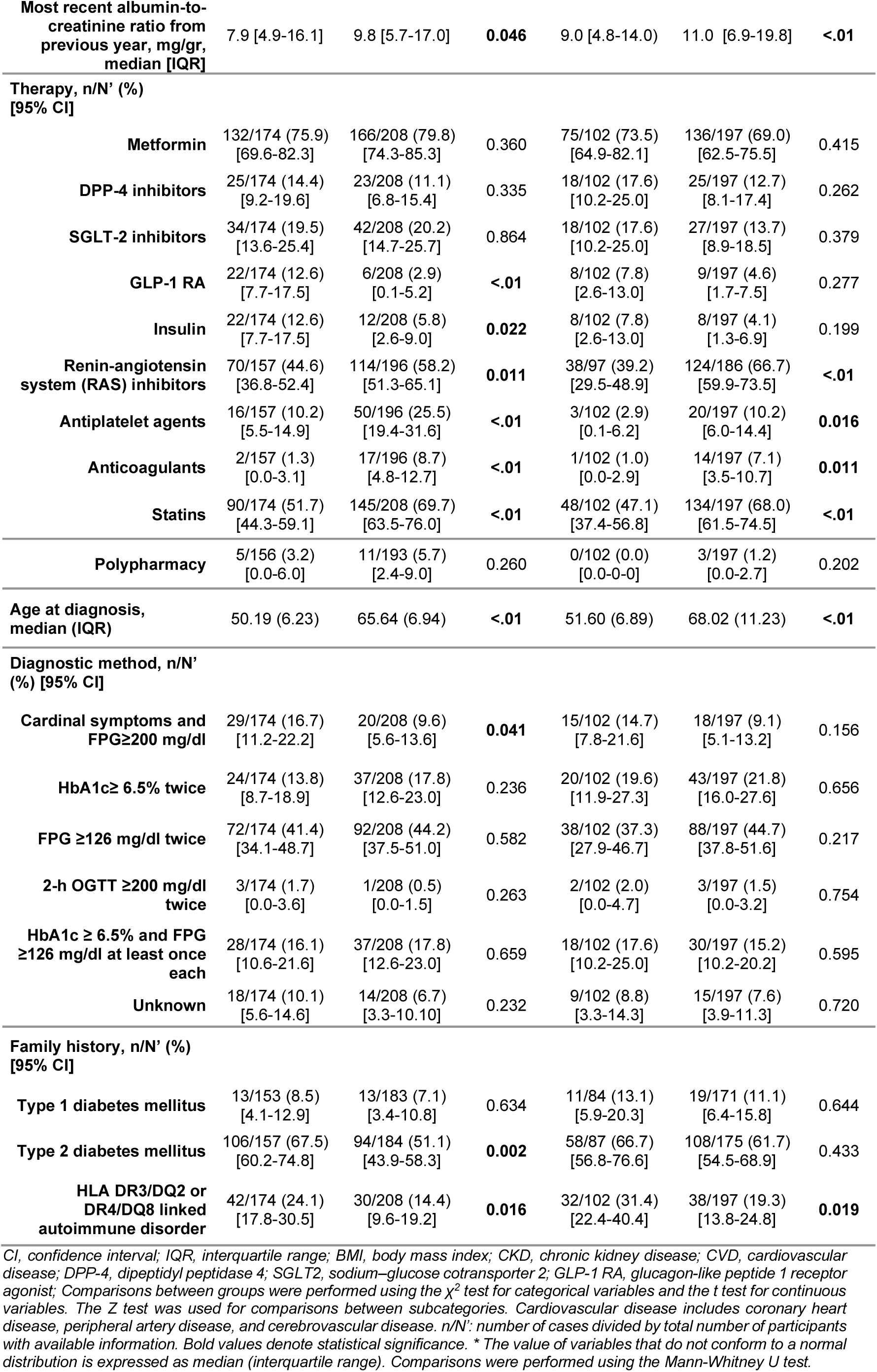
Baseline characteristics by sex and age group.

In terms of physical activity, younger men were significantly more likely to have low activity than older men (52.9% ≤60 vs. 39.9% >60, p=.01). In contrast, older women were more likely to have low levels of physical activity than younger women (41.2% ≤60 vs. 58.4% >60, p<.01). Regarding diet, there were differences by age for men but not for women in terms of consumption of wine (13.8% ≤60 vs. 31.4% >60, p<.01), olive oil (91.3% ≤60 vs. 97.9% >60, p<0.01), fruit (46.6% ≤60 vs. 60.1% >60, p=0.011), and red meat (63.8% ≤60 vs. 75.8% >60, p=0.014). Older men consumed more wine, olive oil and fruit and red meat than younger men (Table 4).

**Table 4.**
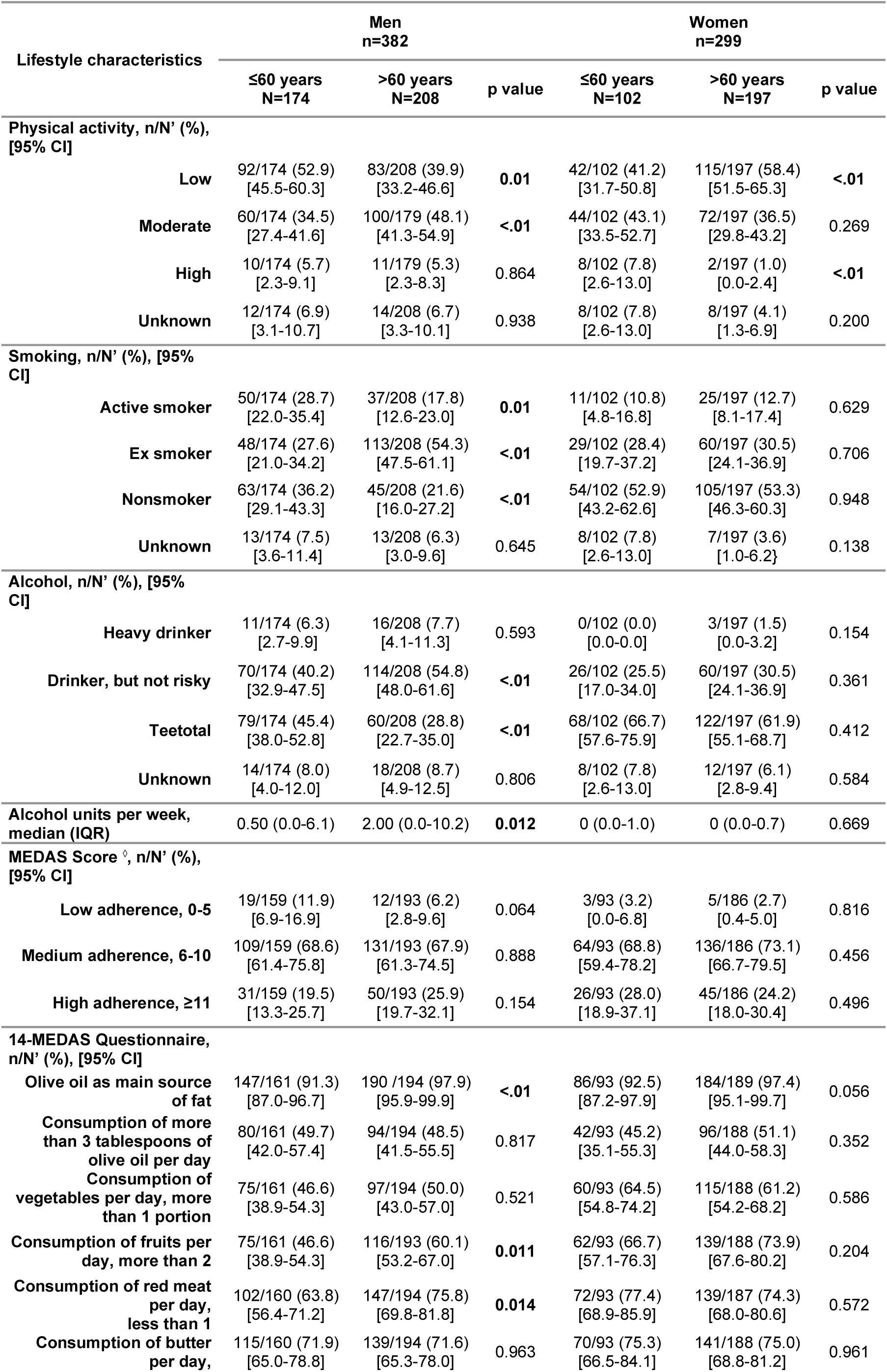
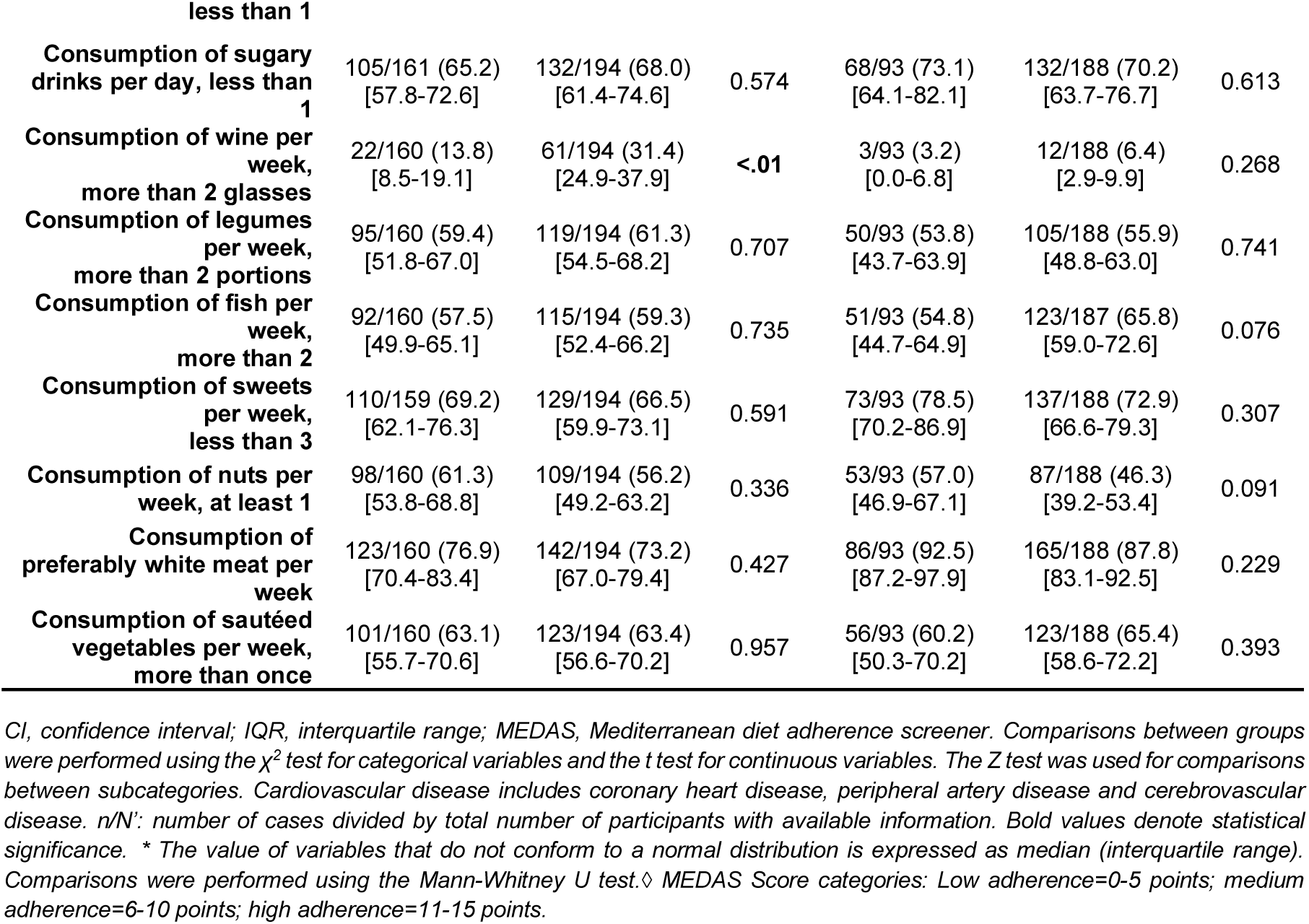
Lifestyle characteristics by sex and age group.

### Patient characteristics by age group and sex

Differences in sex frequencies were observed by age group (see Supplementary Table 1-2). In terms of biochemical parameters, younger men at diagnosis showed higher median FPG values than women (169 [IQR, 133-250] vs. 145 [IQR, 125-193], p<.01), as well as higher median HbA1c values (7.8 [IQR, 6.8-10.3] vs. 7.0 [IQR, 6.4-8.3], p<.01). Similar results were found in older men compared to older women. Additionally, younger men displayed higher median triglyceride levels than younger women (146 [IQR, 100-212] vs. 113 [IQR, 81-156], p <.01), whereas this difference was not observed in older men.

Regarding HDL and LDL cholesterol, both were higher in younger and older women compared to men. Metformin was used more often in men than in women in the older age group (69% women vs. 79.8% men, p=0.013).

Women older than 60 years, more frequently had a family history of T2DM than men of the same age (61.7% women vs. 51.1% men, p=0.042).

As for physical activity, women in the older age group were more frequently less active (58.4% women vs. 39.9% men, p<.001).

Differences in adherence to the Mediterranean diet were observed in the younger age group. Adherence was poor in 3.2% of women and 11.9% of men (p=0.012).

### Association between sex, age, educational level, adherence to the Mediterranean diet, and high physical activity

Multivariate logistic regression analysis showed that patients with a high educational level (OR= 1.90, 95% CI, 1.28-2.81)) and those >60 years (OR= 1.49; 95% CI, 1.01-2.21 were more adherent to the Mediterranean diet, and the older ones were associated with low intensity exercise (OR= 0.34, 95% CI, 0.16-0.75). Normal blood pressure was associated with Mediterranean diet and high physical activity and body mass index was inversely associated with physical activity (Table 5).

**Table 5.** Multivariate logistic regression analysis of the associations between sex, age, and education level and adherence to the Mediterranean diet and high physical activity.

| Adherence to Mediterranean diet |  |  |  | High physical activity |  |  |
| --- | --- | --- | --- | --- | --- | --- |
|  | OR | 95% CI | p value | OR | 95% CI | p value |
| Sex (women) | 1.17 | 0.81-1.70 | 0.401 | 0.46 | 0.20-1.05 | 0.066 |
| Age (>60 years) | 1.49 | 1.01-2.21 | 0.048 | 0.34 | 0.16-0.75 | 0.007 |
| High educational level | 1.90 | 1.28-2.81 | 0.001 | 1.52 | 0.70-3.31 | 0.286 |
| Normal blood pressure (yes vs. no) | 1.52 | 1.05-2.21 | 0.027 | 4.03 | 1.69-9.61 | 0.002 |
| DPP-4 inhibitors current use | 0.66 | 0.41-1.06 | 0.085 |  |  |  |
| DPP-4 inhibitors previous use | 0.22 | 0.03-1.86 | 0.164 | - |  |  |
| BMI (per 1 kg/m2 increase) |  |  |  | 0.92 | 0.85-0.99 | 0.028 |
| Smoking |  |  |  |  |  |  |
| Current smoker |  |  |  | 0.68 | 0.23-2.03 | 0.487 |
| Previous smoker |  |  |  | 1.58 | 0.72-3.47 | 0.259 |
| Alcohol |  |  |  |  |  |  |
| Risky use |  |  |  | 3.88 | 0.88-17.1 | 0.074 |
| Non-risky use |  |  |  | 1.86 | 0.86-3.99 | 0.113 |

## Discussion

The LADA-PC study is a cross-sectional, observational research study performed in Madrid (Spain) and involving adults recently diagnosed with DM. One of its objectives is to describe the sociodemographic and clinical characteristics and health parameters associated of this population.

The most notable finding is the high frequency of multiple cardiovascular risk factors and associated conditions, even in the early stages of the disease. Patients in the LADA-PC study are relatively young (59 years at diagnosis), and about 60% have hypercholesterolemia, hypertension, obesity and meet the criteria for metabolic syndrome; a quarter have hypertriglyceridemia. Despite being recently diagnosed, about 8% have microalbuminuria and 10% have atherosclerotic CVD. The proportion of unhealthy lifestyles found is high, as evidenced by smoking (19%), low physical activity (52%), alcohol consumption (43%) and excess alcohol consumption (5%). More than half have a family history of T2DM.

A Danish study performed in a dynamic cohort of patients recently diagnosed with T2DM (7,011 people evaluated) yielded similar results. The median age at diagnosis was 61 years, 55% of women and 52% of men were obese, 16% had microalbuminuria, 69% were treated for hypertension and 66% for cholesterol, and 53% had a family history of diabetes mellitus. Furthermore, 61% did not exercise regularly, 18% were smokers and 7% were risk drinkers [22].

In another prospective dynamic cohort study of 12,673 people with T2DM enrolled 0.7 years after diagnosis and followed since 1998 in the Netherlands, the mean age was 63 years, 21% smoked, and the annual incidence was 3-4% for CVD, 2.4-9.6% for kidney failure, and , 0.8-1.8% for retinopathy [23].

Multiple studies have shown that healthy lifestyle choices and weight loss can delay metabolic deterioration and the onset of complications in newly diagnosed T2DM [24,25].

A recent study of 7,847 individuals with new-onset T2DM in China [26] found that a combined association of metabolic risk components accounted for 36.1% of the risk attributable to diabetes mellitus and that physical inactivity, unhealthy diet, and smoking were strongly associated with major cardiovascular events.

According to the German Diabetes Study [27], good adherence to a healthy lifestyle compared to poor adherence was associated with lower levels of triglycerides, cholesterol, glycosylated hemoglobin, high-sensitivity C-reactive protein, and hepatic and visceral fat in newly diagnosed T2DM patients.

Similarly, promoting physical activity and avoiding a sedentary lifestyle are essential for preventing and improving control of diabetes mellitus [28,29].

In the LADA-PC population, we found that high adherence to the Mediterranean diet was high in only 24% of patients and medium or low adherence in 69%. We also found sex differences regarding consumption of fruits and vegetables, with lower consumption recorded in men. Consumption of extra virgin olive oil, on the other hand, is almost universal in the whole population, although it is still below the recommended 4 tablespoons. This consumption may decrease in the coming years owing to the significant increase in the price of this product.

One aspect of the Mediterranean diet, consumption of red wine, is controversial, since recommendation of this product to patients may entail a risk of addiction and multiple additional health problems associated with alcohol. Recent studies have also shown that alcohol abstinence in newly diagnosed diabetic adults is associated with a low risk of developing atrial fibrillation [30]. In the LADA-PC study, 43% of patients (53% of men and 31% of women) drank alcohol regularly, and 5% (8% of men and 1% of women) were high-risk drinkers.

The presence of so many comorbidities and risk factors from the onset of diabetes mellitus, reminds us of the importance of promoting health education for society in general and for patients with diabetes mellitus in particular, in addition to intensifying pharmacological treatments that have demonstrated metabolic benefits and reduced morbidity and mortality. Studies show that this approach is essential not only for the lifestyle intervention itself, but also for its quality and individualization, because motivation for change after a diagnosis of diabetes mellitus is complex and can be affected by poor self-determination [31,32,33].

Among the most common diabetes mellitus medications, metformin stands out with prescription to 80% of patients, followed by the group comprising SGLT-2 inhibitors (19%), DPP-4 inhibitors (14%), GLP-1 RA (7%) and insulin (8%). Following the recommendations of clinical guidelines, SGLT-2 inhibitors and GLP-1 RA will be prescribed more frequently owing to their nephroprotective and cardioprotective properties.

In the absence of contraindications, metformin should be initiated as a first-line therapy at disease onset. A recent Taiwanese study of 474,410 patients with T2DM and a mean follow-up of 5.8 years showed that, metformin administered early after diagnosis was associated with fewer major adverse cardiac events compared with patients who did not receive metformin [34].

Regarding other cardiovascular treatments, above 51% of patients in the LADA-PC study received a renin-angiotensin inhibitor for arterial hypertension, sixty-four percent received lipid-lowering treatment and 14% antiplatelet agents.

A recent survey conducted in several European countries [35] showed that a high proportion of patients with diabetes mellitus (patients newly diagnosed with DM and patients with a known history of DM) and coronary heart disease do not receive optimal treatment according to guideline recommendations: 55% of patients newly diagnosed with DM and 60% of patients with known DM did not receive all four cardioprotective medications (statins, beta-blockers, renin-angiotensin system blockers, and antiplatelet agents); 39% of newly diagnosed diabetic patients and 46% of known diabetic patients had poorly controlled blood pressure, and only 18% of newly diagnosed diabetic patients and 28% of known diabetic patients had LDL cholesterol levels <70 mg/dl.

Being a woman and being over 60 years of age were positively associated with adherence to the Mediterranean diet but negatively associated with high physical activity. High education levels and normal blood pressure were linked to adherence to the Mediterranean diet and physical activity. On the other hand, for each unit increase in BMI, the prevalence of high physical activity decreased by 8% (OR= 0.92; 95% CI, 0.85-0.99).

These findings are consistent with those of other studies supporting this association [36–38]. The relationship between high levels of education and a healthy lifestyle has been confirmed in an international study led by scientists at King’s College London, who showed that older adults with a university education and higher incomes enjoy more active and healthy aging than their peers with a lower educational and economic levels [39].

Our findings showed that normal blood pressure was associated with Mediterranean diet. The cardiovascular benefits of this diet have been demonstrated in multiple studies and, in addition, it is possible that participants without hypertension have fewer limitations in physical activity.

However, individuals with a high BMI experience difficulty in practicing physical activity, as highlighted elsewhere [40], although a U-shaped relationship between BMI and physical activity has been described in younger persons [41].

Regarding nutrition, an unhealthy diet could lead to traditional risk factors, such as higher LDL, and triglycerides levels and hypertension, while accelerating the progression of atherosclerosis [42]. It would therefore be reasonable to encourage patients newly diagnosed with DM and with normal blood pressure follow a healthy diet such as the Mediterranean diet to avoid other risk factors such as hypertension and dyslipidemia.

This study has several limitations. The first is inherent to all cross-sectional studies; namely, the lack of directionality precludes the associations found from indicating causality. The second is that given the large number of healthcare professionals involved in patient recruitment, there is no total guarantee that patients were selected in accordance with the protocol and that convenience sampling was not used. Third, as the questionnaires were administered in a healthcare setting, participants’ responses may have been conditioned by a desire to please the professionals who care for them. Fourth, there were losses of nearly 10% for certain variables, so the 95% confidence interval may differ from the calculation if data had been collected from all subjects. Therefore, caution should be taken when inferring to the general population. Finally, the criterion for recent diagnosis included patients diagnosed up to four years earlier, and complications associated with DM might have been less common if the period of recent diagnosis had been shorter.

On the other hand, this study has several strengths. The first is its large sample of patients newly diagnosed with DM, which enables stratification by sex and age with sufficient statistical power to detect differences, if any. Second, the use of a centralized blood sampling laboratory reduces the likelihood of bias in the diagnostic classification of DM, especially if different analytical techniques have been used. Finally, since patients were recruited from multiple health centers and had a wide range of socioeconomic levels, the study population is a representative sample of all patients newly diagnosed with DM in the Community of Madrid.

In conclusion, sex differences were found, with morbid obesity, and poor lipid profile in women, and treatment with metformin and antiplatelet agents were less frequent in women than in men. However, they had fewer comorbidities than men. Similarly, the >60 years subgroups of men and women had higher burdens of vascular disease (CVD, atrial fibrillation), hypertension, and dyslipidemia. Older groups achieved better lipid and glycemic control, the first, most likely related to more frequent use of statins.

In general, patients newly diagnosed with DM in the population of Madrid are characterized by major cardiovascular risk factors and comorbidities in the early stages of the disease. Many of these conditions can be modified through a healthy lifestyle and weight reduction. Raising awareness of this significant problem among patients and professionals will improve the population’s health, prevent cardiovascular events, and reduce mortality.

## Data Availability

There are restrictions on data availability for the LADA-AP study due to the signed
consent agreements around data sharing, which only allow access to external
researchers for research following the project's purposes once it has been completed.
Requestors wishing to access the LADA-AP data used in this study can request it to
the LADA-AP Steering Committee.

## Additional information

### Author Contributions

**Conceptualization**: Vich-Pérez P, Salinero-Fort MA

**Data curation:** Taulero-Escalera B, Salinero-Fort MA.

**Formal analysis:** Salinero-Fort MA, Taulero-Escalera B.

**Funding acquisition**: Vich-Pérez P.

**Investigation**: Vich-Pérez P, García-Espinosa V, Villanova-Cuadra L, Regueiro-Toribio P, Sevilla-Machuca I, Timoner-Aguilera J, Martínez-Grandmontagne M, Abós-Pueyo T, Álvarez-Hernandez-Cañizares C, Reviriego-Jaén G, Serrano-López-Hazas A, Gala-Molina I, Sanz-Pascual M.

**Methodology**: Salinero-Fort MA, Taulero-Escalera B,

**Project administration**: Salinero-Fort MA.

**Supervision**: Salinero-Fort MA, Taulero-Escalera B, Vich-Pérez P.

**Validation**: Vich-Pérez P, Taulero-Escalera B, Villanova-Cuadra L, Timoner-Aguilera J, Martínez-Grandmontagne M, Álvarez-Hernandez-Cañizares C, Reviriego-Jaén G, Serrano-López-Hazas A, Gala-Molina I, Sanz-Pascual M.

**Writing – original draft:** Salinero-Fort MA, Vich-Pérez P, Taulero-Escalera B.

**Writing – Review & Editing:** Taulero-Escalera B, Salinero-Fort MA, Vich-Pérez P, García-Espinosa V, Salinero-Fort MA, Abós-Pueyo T, Regueiro-Toribio P, Sevilla-Machuca I.

## Acknowledgments

**LADA-PC Research Consortium**: Mencia-Valle C (2a), Berzal-Rosende M (2a), Díaz-Sánchez J (8a), Fernández-Alonso N (8b), Camarero-Shelly M (8b), Pérez González F (9a), Gutiérrez-López S (9b), Ortiz I (9b), Bentata-Levy LV (9b), Domínguez-Agüero N (7a), Redondo-Sendino A (7a), Rodríguez-Alonso L (7b), Redondo-Gómez A (7b), Leyva-Vera G (7b), Romero-Molina ML (7b), Toro-Herrero A (7b), Lasheras-Garcia J (7b), Tormo-Ortiz I (10a), Ocaña-Dominguez AM (10a), Sánchez-Fernández R (10a), Luis-García M (10b), Careaga-González I (10b), Fuster-Tozer M (11a), Fernández-López E (11a), Castaño-Rodríguez CA (11a), Santos-Álvarez C (4a), Galán-Gutiérrez MT (4a), Ruiz-Giardín RM (4a), Gracia-Moliner MO (4a), Alonso-Leonardo A (4b), Cuenca-Blanco MT (4b), Ferrero-García MG (4b), Goicoechea-García M (4a), Cárdenas-De Miguel A (1a), Gallego-Fernández J (1a), Puerto-Rodríguez M (1a), Vicente-Mata B (1a), Sánchez-López O (1a), Brusint-Olivares B (1a), Prieto-Checa I (1a), Moreno-Gómez AI (1a), Pérez-Medina SA (1a), Settanni-Gomis C (1a), Moratinos-Recuenco I (1a), Guereña-Tomás MJ (1a), Campos-Campos N, Coleto-Gutierrez R (1a), Madero-Velázquez S (1a), Ayuso-Gil A (1b), Castro-Benito M (1b), Botanes-Peñafiel L (1b), Duque-Rebollo I (1b), García-Ortega AM (1b), Alonso-Roca R (6a), Herrero-Delgado M (6a), Asenjo-Calvo M (6a), Pérez-Medina A (6a), Sánchez-Villares-Rodríguez M (6a), Villar-Coloma E (6a), Criado-Jorge S (6b), García-Campo V (6b), López-Herrera AM (6b), Mallavibarrena-Ramírez B (6b), Muñoz-Millán E (6b), Vargas-Reyes ML(6b), Garcia-Del Río I (6b), Abánades-Herranz JC (3a), Nieto-Gualda M (3b), Pertierra-Galindo N (3a), Muñoz-Quirós-Aliaga S (3a), Domínguez-Moreno E (12a), López-Carrasco M (12b). The adittional members of the LADA-PC Consortium are Roy-Ariño G(13c), Ortega-Sánchez S(13e), Carrasco-Sayalero AM (13c), Iriarte-Campo V (14d), Estévez-Muñoz JC (15a), del Cura-González I (16a), Sanz-Cuesta T (16a), Sanz-del Oso J(15f).

Lead author for this group: Guereña-Tomás MJ (1a).

(a) Family doctor, (b) Nurse, c) Specialist in clinical immunology, (d) Medical researcher, (e) Laboratory technician, (f) Telecommunications engineer (1) Los Alpes Health Center, (2) Aquitania Health Center, (3) Monóvar Health Center, (4) García Noblejas Health Center, (5) Alameda de Osuna Health Center, (6) Mar Baltico Health Center, (7) Canillejas Health Center, (8) Barajas Health Center, (9) Benita de Ávila Health Center, (10) Dr Cirajas Health Center, (11) Estrecho de Corea Health Center, (12) Sanchinarro Health Center, (13) Ramón y Cajal Hospital. (14) FIIBAP, (15)SERMAS Primary Care, (16) Research Unit Primary Care Management.

## Notes

### Competing Interest Statement

The authors have declared no competing interest.

### Clinical Protocols

https://journals.plos.org/plosone/article/authors?id=10.1371/journal.pone.0281657

### Funding Statement

This study has been funded by Instituto de Salud Carlos III (ISCIII) through the project
"PI19/01569" and co-funded by the European Union. The funders had no role in study
design, data collection and analysis, decision to publish, or preparation of the
manuscript

### Author Declarations

Research and Medicines Ethics Committee and the Central Research Commission of the Primary Care Management of Madrid gave ethical approval for this work, with reference number: CODE: JCAH/PVP/LVP/LVP/LVP/LVP/LADA/2019/1. VERSION: March 4, 2019.

## References

1. Roden M, Shulman GI. The integrative biology of type 2 diabetes. Nature. diciembre de 2019;576(7785):51-60.

2. Soriguer F, Goday A, Bosch-Comas A, Bordiú E, Calle-Pascual A, Carmena R, et al. Prevalence of diabetes mellitus and impaired glucose regulation in Spain: the Study. Diabetologia. enero de 2012;55(1):88–93.

3. Diabetes mellitus. Report of a WHO Study Group. World Health Organ Tech Rep Ser. 1985;727:1–113.

4. Alberti KG, Zimmet PZ. Definition, diagnosis and classification of diabetes mellitus and its complications. Part 1: diagnosis and classification of diabetes mellitus provisional report of a WHO consultation. Diabet Med J Br Diabet Assoc. julio de 1998;15(7):539–53.

5. Tuomilehto J, Lindström J, Eriksson JG, Valle TT, Hämäläinen H, Ilanne-Parikka P, et al. Prevention of type 2 diabetes mellitus by changes in lifestyle among subjects with impaired glucose tolerance. N Engl J Med. 3 de mayo de 2001;344(18):1343–50.

6. Gong Q, Zhang P, Wang J, Gregg EW, Cheng YJ, Li G, et al. Efficacy of lifestyle intervention in adults with impaired glucose tolerance with and without impaired fasting plasma glucose: A post hoc analysis of Da Qing Diabetes Prevention Outcome Study. Diabetes Obes Metab. octubre de 2021;23(10):2385–94.

7. Standards of Medical Care in Diabetes-2016: Summary of Revisions. Diabetes Care. enero de 2016;39 Suppl 1:S4–5.

8. Becerra-Tomás N, Blanco Mejía S, Viguiliouk E, Khan T, Kendall CWC, Kahleova H, et al. Mediterranean diet, cardiovascular disease and mortality in diabetes: A systematic review and meta-analysis of prospective cohort studies and randomized clinical trials. Crit Rev Food Sci Nutr. 2020;60(7):1207–27.

9. Guasch-Ferré M, Willett WC. The Mediterranean diet and health: a comprehensive overview. J Intern Med. septiembre de 2021;290(3):549–66.

10. Schwingshackl L, Missbach B, König J, Hoffmann G. Adherence to a Mediterranean diet and risk of diabetes: a systematic review and meta-analysis. Public Health Nutr. mayo de 2015;18(7):1292–9.

11. Basterra-Gortari FJ, Ruiz-Canela M, Martínez-González MA, Babio N, Sorlí JV, Fito M, et al. Effects of a Mediterranean Eating Plan on the Need for Glucose-Lowering Medications in Participants With Type 2 Diabetes: A Subgroup Analysis of the PREDIMED Trial. Diabetes Care. agosto de 2019;42(8):1390–7.

12. Colberg SR, Sigal RJ, Yardley JE, Riddell MC, Dunstan DW, Dempsey PC, et al. Physical Activity/Exercise and Diabetes: A Position Statement of the American Diabetes Association. Diabetes Care. noviembre de 2016;39(11):2065–79.

13. American Diabetes Association. 5. Facilitating Behavior Change and Well-being to Improve Health Outcomes: Standards of Medical Care in Diabetes-2020. Diabetes Care. enero de 2020;43(Suppl 1):S48–65.

14. Pérez Unanua MP, Alonso Fernández M, López Simarro F, Soriano Llora T, Peral Martínez I, Mancera Romero J, et al. [Adherence to healthy lifestyle behaviours in patients with type 2 diabetes in Spain]. Semergen. abril de 2021;47(3):161–9.

15. Schlesinger S, Neuenschwander M, Ballon A, Nöthlings U, Barbaresko J. Adherence to healthy lifestyles and incidence of diabetes and mortality among individuals with diabetes: a systematic review and meta-analysis of prospective studies. J Epidemiol Community Health. mayo de 2020;74(5):481–7.

16. Vich-Pérez P, Abánades-Herranz JC, Mora-Navarro G, Carrasco-Sayalero ÁM, Salinero-Fort MÁ, Sevilla-Machuca I, et al. Development and validation of a clinical score for identifying patients with high risk of latent autoimmune adult diabetes (LADA): The LADA primary care-protocol study. PloS One. 2023;18(2):e0281657.

17. American Diabetes Association. 2. Classification and Diagnosis of Diabetes: Standards of Medical Care in Diabetes-2019. Diabetes Care. enero de 2019;42(Suppl 1):S13–28.

18. National Cholesterol Education Program (NCEP) Expert Panel on Detection, Evaluation, and Treatment of High Blood Cholesterol in Adults (Adult Treatment Panel III). Third Report of the National Cholesterol Education Program (NCEP) Expert Panel on Detection, Evaluation, and Treatment of High Blood Cholesterol in Adults (Adult Treatment Panel III) final report. Circulation. 17 de diciembre de 2002;106(25):3143–421.

19. Crespo-Salgado JJ, Delgado-Martín JL, Blanco-Iglesias O, Aldecoa-Landesa S. [Basic guidelines for detecting sedentarism and recommendations for physical activity in primary care]. Aten Primaria. marzo de 2015;47(3):175–83.

20. Schröder H, Fitó M, Estruch R, Martínez-González MA, Corella D, Salas-Salvadó J, et al. A short screener is valid for assessing Mediterranean diet adherence among older Spanish men and women. J Nutr. junio de 2011;141(6):1140–5.

21. Masnoon N, Shakib S, Kalisch-Ellett L, Caughey GE. What is polypharmacy? A systematic review of definitions. BMC Geriatr. 10 de octubre de 2017;17(1):230.

22. Christensen DH, Nicolaisen SK, Berencsi K, Beck-Nielsen H, Rungby J, Friborg S, et al. Danish Centre for Strategic Research in Type 2 Diabetes (DD2) project cohort of newly diagnosed patients with type 2 diabetes: a cohort profile. BMJ Open. 7 de abril de 2018;8(4):e017273.

23. van der Heijden AA, Rauh SP, Dekker JM, Beulens JW, Elders P, ’t Hart LM, et al. The Hoorn Diabetes Care System (DCS) cohort. A prospective cohort of persons with type 2 diabetes treated in primary care in the Netherlands. BMJ Open. 6 de junio de 2017;7(5):e015599.

24. Burch E, Williams LT, Thalib L, Ball L. Short-term improvements in diet quality in people newly diagnosed with type 2 diabetes are associated with smoking status, physical activity and body mass index: the 3D case series study. Nutr Diabetes. 13 de julio de 2020;10(1):25.

25. Sarathi V, Kolly A, Chaithanya HB, Dwarakanath CS. High rates of diabetes reversal in newly diagnosed Asian Indian young adults with type 2 diabetes mellitus with intensive lifestyle therapy. J Nat Sci Biol Med. 2017;8(1):60–3.

26. Li M, Xu Y, Wan Q, Shen F, Xu M, Zhao Z, et al. Individual and Combined Associations of Modifiable Lifestyle and Metabolic Health Status With New-Onset Diabetes and Major Cardiovascular Events: The China Cardiometabolic Disease and Cancer Cohort (4C) Study. Diabetes Care. agosto de 2020;43(8):1929–36.

27. Baechle C, Lang A, Strassburger K, Kuss O, Burkart V, Szendroedi J, et al. Association of a lifestyle score with cardiometabolic markers among individuals with diabetes: a cross-sectional study. BMJ Open Diabetes Res Care. julio de 2023;11(4):e003469.

28. Falconer CL, Cooper AR, Walhin JP, Thompson D, Page AS, Peters TJ, et al. Sedentary time and markers of inflammation in people with newly diagnosed type 2 diabetes. Nutr Metab Cardiovasc Dis NMCD. septiembre de 2014;24(9):956–62.

29. Llavero-Valero M, Escalada San Martín J, Martínez-González MA, Alvarez-Mon MA, Alvarez-Alvarez I, Martínez-González J, et al. Promoting exercise, reducing sedentarism or both for diabetes prevention: The «Seguimiento Universidad De Navarra» (SUN) cohort. Nutr Metab Cardiovasc Dis NMCD. 8 de febrero de 2021;31(2):411-9.

30. Choi YJ, Han KD, Choi EK, Jung JH, Lee SR, Oh S, et al. Alcohol Abstinence and the Risk of Atrial Fibrillation in Patients With Newly Diagnosed Type 2 Diabetes Mellitus: A Nationwide Population-Based Study. Diabetes Care. junio de 2021;44(6):1393–401.

31. Sebire SJ, Toumpakari Z, Turner KM, Cooper AR, Page AS, Malpass A, et al. «I’ve made this my lifestyle now»: a prospective qualitative study of motivation for lifestyle change among people with newly diagnosed type two diabetes mellitus. BMC Public Health. 31 de enero de 2018;18(1):204.

32. de Hoogh IM, Pasman WJ, Boorsma A, van Ommen B, Wopereis S. Effects of a 13-Week Personalized Lifestyle Intervention Based on the Diabetes Subtype for People with Newly Diagnosed Type 2 Diabetes. Biomedicines. 10 de marzo de 2022;10(3):643.

33. Pikkemaat M, Boström KB, Strandberg EL. «I have got diabetes!» -interviews of patients newly diagnosed with type 2 diabetes. BMC Endocr Disord. 24 de mayo de 2019;19(1):53.

34. Lee KT, Yeh YH, Chang SH, See LC, Lee CH, Wu LS, et al. Metformin is associated with fewer major adverse cardiac events among patients with a new diagnosis of type 2 diabetes mellitus: A propensity score-matched nationwide study. Medicine (Baltimore). julio de 2017;96(28):e7507.

35. Gyberg V, De Bacquer D, De Backer G, Jennings C, Kotseva K, Mellbin L, et al. Patients with coronary artery disease and diabetes need improved management: a report from the EUROASPIRE IV survey: a registry from the EuroObservational Research Programme of the European Society of Cardiology. Cardiovasc Diabetol. 1 de octubre de 2015;14:133.

36. Mendonça N, Gregório MJ, Salvador C, Henriques AR, Canhão H, Rodrigues AM. Low Adherence to the Mediterranean Diet Is Associated with Poor Socioeconomic Status and Younger Age: A Cross-Sectional Analysis of the EpiDoC Cohort. Nutrients. 15 de marzo de 2022;14(6):1239.

37. Poklar Vatovec T, Jenko Pražnikar Z, Petelin A. Adherence and Sociodemographic Determinants of Adherence to the Mediterranean Diet among Slovenian Adults and the Elderly. Nutrients. 20 de julio de 2023;15(14):3219.

38. Sánchez-Hernández MS, Rodríguez-Caldero MC, Martín-Pérez MP, Mira-Solves JJ, Vitaller-Burillo J, Carratalá-Munuera MC. Impact of adherence to Mediterranean diet and/or drug treatment on glycaemic control in type 2 diabetes mellitus patients: DM2-CUMCYL study. Prim Care Diabetes. diciembre de 2020;14(6):685–91.

39. Wu YT, Daskalopoulou C, Muniz Terrera G, Sanchez Niubo A, Rodríguez-Artalejo F, Ayuso-Mateos JL, et al. Education and wealth inequalities in healthy ageing in eight harmonised cohorts in the ATHLOS consortium: a population-based study. Lancet Public Health. julio de 2020;5(7):e386–94.

40. Cárdenas Fuentes G, Bawaked RA, Martínez González MÁ, Corella D, Subirana Cachinero I, Salas-Salvadó J, et al. Association of physical activity with body mass index, waist circumference and incidence of obesity in older adults. Eur J Public Health. 1 de octubre de 2018;28(5):944-50.

41. Chen G, Chen J, Liu J, Hu Y, Liu Y. Relationship between body mass index and physical fitness of children and adolescents in Xinjiang, China: a cross-sectional study. BMC Public Health. 5 de septiembre de 2022;22(1):1680.

42. Lu L, Jing W, Qian W, Fan L, Cheng J. Association between dietary patterns and cardiovascular diseases: A review. Curr Probl Cardiol. 24 de enero de 2024;49(3):102412.

