## Supplementary Material for "Sex and age differences in cardiovascular risk factors and lifestyle at the onset of diabetes mellitus: a cross-sectional study in Spanish Primary Health Care"

**S1 Table. Baseline characteristics by age group and sex.**

| Baseline characteristics | ≤60 years  N= 276 | | | >60 years  N= 405 | | |
| --- | --- | --- | --- | --- | --- | --- |
|  | **Men**  **N=174**  **(63%)** | **Women**  **N=102**  **(37%)** | **p value** | **Men**  **N=208**  **(51.4%)** | **Women**  **N=197**  **(48.6%)** | **p value** |
| Education, n (%) [95% CI] |  |  |  |  |  |  |
| Illiterate or incomplete primary education | 1 (0.6) [0.0-1.8] | 2 (2.0)  [0.0-4.7] | 0.387 | 10 (4.8) [1.9-7.7] | 29 (14.7) [9.8-19.6] | **<.001** |
| Primary education | 13 (7.5) [3.6-11.4] | 5 (4.9) [0.1-9.1] | 0.569 | 55 (26.4) [20.4-32.4] | 69 (35.0) [28.3-41.7] | 0.061 |
| Elementary High School, | 31 (17.8) [12.1-23.5] | 20 (19.6) [11.9-27.3] | 0.711 | 40 (19.2) [13.9-24.6] | 38 (19.3) [13.8-24.8] | 0.980 |
| Industrial Officer's degree | 17 (9.8) [5.4-14.2] | 17 (16.7) [9.5-23.9] | 0.103 | 13 (6.3) [3.0-9.6] | 10 (5.1) [2.0-8.2] | 0.603 |
| Superior High School | 24 (13.8) [8.7-18.9] | 15 (14.7) [7.8-21.6] | 0.836 | 26 (12.5) [8.0-17.0] | 24 (12.2) [7.6-16.8] | 0.927 |
| Industrial Master's Degree | 33 (19.0) [13.2-24.8] | 13 (12.7) [6.2-19.2] | 0.167 | 22 (10.6) [6.4-14.8] | 11 (5.6) [2.4-8.8] | 0.065 |
| University | 51 (29.3) [22.5-36.1] | 25 (24.5) [16.2-32.9] | 0.385 | 37 (17.8) [12.6-23.0] | 14 (7.1) [3.5-10.7] | **<.001** |
| Unknown | 4 (2.3) [0.1-4.5] | 5 (4.9) [0.7-9.1] | 0.263 | 5 (2.4) [0.3-4.5] | 2 (1.0) [0.0-2.4] | 0.276 |
| BMI, kg/m^2^, median [IQR] | 31.1 [27.8-34.5] | 30.2 [25.5-33.7] | 0.471 | 29.6 [26.8-32.4] | 29.6 [26.1-33.7] | 0.971 |
| BMI categories, n/N’ (%) [95% CI] |  |  |  |  |  |  |
| <25 kg/m^2^ | 14/163 (8.6) [4.3-12.9] | 18/92 (19.6) [11.5-27.7] | **0.015** | 20/188 (10.6) [6.2-15.0] | 35/185 (18.9) [13.3-24.5] | **0.024** |
| 25-29 kg/m^2^ | 41/163 (25.2) [18.5-31.9] | 23/92 (25.0) [16.2-33.9] | 0.973 | 58/188 (30.9) [24.3-37.5] | 49/185 (26.5) [20.1-32.9] | 0.348 |
| 30-34 kg/m^2^ | 71/163 (43.6) [36.0-51.2] | 31/92 (33.7) [24.0-43.4] | 0.132 | 85/188 (45.2) [38.1-52.3] | 61/185 (33.0) [26.2-39.8] | **0.016** |
| 35-40 kg/m^2^ | 25/163 (15.3) [9.8-20.8] | 9/92 (9.8) [3.7-15.9] | 0.219 | 21/188 (11.2) [6.7-15.7] | 24/185 (13.0) [8.2-17.9] | 0.594 |
| >40 kg/m^2^ | 12/163 (7.4) [3.4-11.4] | 11/92 (12.0) [5.4-18.6] | 0.233 | 4/188 (2.1) [0.5-4.2] | 16/185 (8.6) [4.6-12.6] | **<.01** |
| Waist circumference, cm, median [IQR] | 109 [100-119] | 101 [90-113) | **<.01** | 108 [101-114] | 102 [93-112] | **<.001** |
| Comorbidities, n/N’ (%) [95% CI] |  |  |  |  |  |  |
| Microalbuminuria | 13/140 (9.3) [4.5-14.1] | 4/80 (5.0) [0,2-9.8] | 0.252 | 19/167 (11.4) [6.6-16.2] | 10/167 (6.0) [2.4-9.6] | 0.080 |
| CKD | 2/154 (1.3) [0.0-3.1] | 2/92 (2.2) [0.1-5.2] | 0.599 | 11/188 (5.9) [2.5-9.3] | 8/180 (4.4) [1.4-7.4] | 0.542 |
| Retinopathy | 3/139 (2.2) [0.1-4.6] | 0/86 (0.0) [0.0-0.0] | 0.170 | 5/166 (3.0) [0.4-5.6] | 1/163 (0.6) [0.0-1.8] | 0.104 |
| Neuropathy | 4/149 (2.7) [0.1-5.3] | 1/89 (1.1) [0.0-3.3] | 0.417 | 2/186 (1.1) [0.0-2.6] | 2/174 (1.1) [0.0-2.7] | 0.947 |
| Hypertension | 67/154 (43.5) [35.7-51.3] | 33/94 (35.1) [25.5-44.8] | 0.191 | 118/193 (61.1) [54.2-68.0] | 125/180 (69.4) [62.7-76.1] | 0.093 |
| Hypercholesterolemia | 86/154 (55.8) [48.0-63.6] | 49/93 (52.7) [42.6-62.9] | 0.629 | 135/194 (69.6) [63.1-76.1] | 130/180 (72.2) [65.7-78.8] | 0.575 |
| Hypertriglyceridemia | 49/154 (31.8) [24.4-39.2] | 25/93 (26.9) [17.9-35.9] | 0.412 | 44/190 (23.2) [17.2-29.2] | 38/180 (21.1) [15.1-27.1] | 0.636 |
| Metabolic syndrome | 96/153 (62.3) [54.6-70.0] | 49/90 (54.4) [44.1-64.7] | 0.228 | 97/182 (53.3) [46.1-60.6] | 108/173 (62.4) [55.2-69.6] | 0.083 |
| CVD | 12/174 (6.9) [3.1-10.7] | 2/102 (2.0) [0.0-4.7] | 0.057 | 42/208 (20.2) [14.7-25.7] | 14/197 (7.1) [3.5-10.7] | **<.01** |
| Heart failure | 2/156 (1.3) [0.0-3.1] | 0/93 (0.0) [0.0-0.0] | 0.273 | 4/189 (2.1) [0.1-4.1] | 4/179 (2.2) [0.1-4.4] | 0.938 |
| Atrial fibrillation | 2/158 (1.3) [0.0-3.1] | 1/94 (1.1) [0.0-3.2] | 0.886 | 11/190 (5.8) [2.5-9.1] | 11/180 (6.1) [2.6-9.6] | 0.896 |
| Biochemical parameters |  |  |  |  |  |  |
| FPG at diagnosis, mg/dl, median [IQR] | 169 [133-250] | 145 [125-193] | **<.01** | 138 [127-177] | 133 [123-148] | **<.01** |
| Hb A1c at diagnosis, %, median [IQR] | 7.8 [6.8-10.3] | 7.0 [6.4-8.3] | **<.01** | 7.0 [6.5-7.9] | 6.8 [6.5-7.2] | **0.023** |
| Hb A1c most recent in last 6 months, %, median [IQR] | 6.7 [6.0-7.3] | 6.2 [5.9-6.8] | **0.010** | 6.4 [6.1-6.9] | 6.5 [6.1-6.9] | 0.574 |
| Total cholesterol most recent in last year, mg/dl, median [IQR] | 169  [152-195] | 184  [153-205] | 0.066 | 160 [138-187] | 182 [160-207] | **<.001** |
| HDL cholesterol most recent in last year, mg/dL, median [IQR] | 42 [35-47] | 51 [45-58] | **<.001** | 45 [38-51] | 54 [47-61] | **<.001** |
| LDL cholesterol most recent in last year, mg/dl, mean (SE) | 99 (2.8) | 111 (3.9) | **0.017** | 91 (2.5) | 102 (2.5) | **<.01** |
| Triglycerides most recent in last year, mg/dl, median [IQR] | 146 [100-212] | 113 [81-156] | **<.01** | 115 [87-148] | 114 [93-158] | 0.661 |
| Albumin-to-creatinine ratio most recent in last year, mg/gr* | 8 [5-16) | 9 [5-14] | 0.966 | 10 [6-17] | 11 [7-20] | 0.166 |
| Use of therapies, n/N’ (%) [95% CI] |  |  |  |  |  |  |
| Metformin | 132/174(75.9) [69.6-82.3] | 75/102 (73.5) [64.9-82.1] | 0.658 | 166/208 (79.8) [74.3-85.3] | 136/197 (69.0) [62.5-75.5] | **0.013** |
| iDPP-4 | 25/174 (14.4) [9.2-19.6] | 18/102 (17.6) [10.2-25.0] | 0.484 | 23/208 (11.1) [6.8-15.4] | 25/197 (12.7) [8.1-17.4] | 0.619 |
| iSGLT-2 | 34/174 (19.5) [13.6-25.4] | 18/102 (17.6) [10.2-25.0] | 0.695 | 42/208 (20.2) [14.7-25.7] | 27/197 (13.7) [8.9-18.5] | 0.081 |
| GLP-1 RA | 22/174 (12.6) [7.7-17.5] | 8/102 (7.8) [2.6-13.0] | 0.203 | 6/208 (2.9) [0.6-5.2] | 9/197 (4.6) [1.7-7.5] | 0.368 |
| Insulin | 22/174 (12.6) [7.7-17.5] | 8/102 (7.8) [2.6-13.0] | 0.203 | 12/208 (5.8) [2.6-9.0] | 8/197 (4.1) [1.3-6.9] | 0.435 |
| Antihypertensives therapies | 70/157 (44.6) [36.8-52.4] | 38/97 (39.2) [29.5-48.9] | 0.397 | 114/196 (58.2) [51.3-65.1] | 124/186 (66.7) [59.9-73.5] | 0.087 |
| Antiplatelet | 16/157 (10.2) [5.5-14.9] | 3/97 (3.1) [0.0-6.6] | **0.027** | 50/197 (25.4) [19.3-31.5] | 20/186 (10.8) [6.3-15.3] | **<.001** |
| Anticoagulants | 2/157 (1.3) [0.0-3.1] | 1/97 (1.0) [0.0-3.0] | 0.828 | 17/197 (8.6) [4.7-12.5] | 14/187 (7.5) [3.7-11.3] | 0.692 |
| Statins | 90/165 (54.5) [46.9-62.1] | 48/98 (49.00) [39.1-58.9] | 0.388 | 145/199 (72.9) [66.7-79.1] | 134/189 (70.9) [64.4-77.4] | 0.661 |
| Polypharmacy | 5/156 (3.2) [0.1-6.0] | 0/96 (0.0) [0.0-0.0] | 0.061 | 11/193 (5.7) [0.2-9.0] | 3/180 (1.7) [0.0-3.6] | **0.041** |
| Age at onset of diagnosis, median [IQR] | 51 [46-54) | 51 [48-55] | 0.213 | 64 [60-69] | 68 [61-75] | **<.001** |
| Diagnostic method, n/N’ (%) [95% CI] |  |  |  |  |  |  |
| Cardinal symptoms and glycemia ≥200 mg/dl | 29/174 (16.7) [11.2-22.2] | 15/102 (14.7) [7.8-21.6] | 0.659 | 20/208 (9.6) [5.6-13.6] | 18/197 (9.1) [5.1-13.1] | 0.863 |
| Hb A1c≥ 6.5% twice | 24/174 (13.8) [8.7-18.9] | 20/102 (19.6) [11.9-27.3] | 0.212 | 37/208 (17.8) [12.6-23.0] | 43/197 (21.8) [16.0-27.6] | 0.313 |
| FPG ≥126 mg/dl twice | 72/174 (41.4) [34.1-48.7] | 38/102 (37.3) [27.9-46.7] | 0.501 | 92/208 (44.2) [37.5-51.0] | 88/197 (44.7) [37.8-51.6] | 0.914 |
| 2-h OGTT ≥200 mg/dl twice | 3/174 (1.7) [0.0-3.6] | 2/102 (2.0) [0.0-4.7] | 0.858 | 1/208 (0.5) [0.0-1.5] | 3/197 (1.5) [0.0-3.2] | 0.312 |
| Hb A1c ≥ 6.5% and FPG ≥126 mg/dl at least once each | 28/174 (16.1) [10.6-21.6] | 18/102 (17.6) [10.2-25.0] | 0.748 | 44/208 (21.2) [15.7-26.8] | 30/197 (15.2) [10.2-20.2] | 0.118 |
| Unknown | 12/174 (6.9) [3.1-10.7] | 3/102 (2.9) [0.0-6.2] | 0.137 | 8/208 (3.8) [1.2-6.4] | 8/197 (4.1) [1.3-6.9] | 0.877 |
| Family History, n/N’ (%) [95% CI] |  |  |  |  |  |  |
| Type 1 Diabetes Mellitus | 13/153 (8.5) [4.1-12.9] | 11/84 (13.1) [5.9-20.3] | 0.262 | 13/183 (7.1) [3.4-10.8] | 19/171 (11.1) [6.4-15.8] | 0.189 |
| Type 2 Diabetes Mellitus | 106/157 (67.5) [60.2-74.8] | 58/87 (66.7) [56.8-76.6] | 0.892 | 94/184 (51.1) [43.9-58.3] | 108/175 (61.7) [54.5-68.9] | **0.042** |
| HLA DR3/DQ2 or DR4/DQ8 linked autoimmune disorder | 42/174 (24.1) [17.8-30.5] | 32/102 (31.4) [22.4-40.4] | 0.190 | 30/190 (14.4) [9.4-19.4] | 38/197 (19.3) [13.8-24.8] | 0.190 |

*FPG, fasting plasma glucose; Comparisons between groups were performed using the χ2 test for categorical variables and the t test for continuous variables. The Z test was used for comparisons between subcategories. Cardiovascular disease considers coronary heart disease, peripheral artery disease and cerebrovascular disease. n/N’: number of cases divided by total number of participants with available information. Bold values denote statistical significance.* ** The measure of variables that do not conform to a normal distribution is given in median (interquartile range) and comparison were performed using Mann-Whitney U test.*

**S2 Table. Lifestyle characteristics by age group and sex.**

| Lifestyle characteristics | ≤60 years  N=276 | | | >60 years  N=405 | |  |
| --- | --- | --- | --- | --- | --- | --- |
|  | **Men**  **N=174**  **(63%)** | **Women**  **N=102**  **(37%)** | **p value** | **Men**  **N=208**  **(51.4%)** | **Women**  **N=197**  **(48.6%)** | **p value** |
| Physical activity, n (%) |  |  |  |  |  |  |
| Low | 92 (52.9) | 42 (41.2) | 0.060 | 83 (39.9) | 115 (58.4) | **<.001** |
| Moderate | 60 (34.5) | 44 (43.1) | 0.157 | 100 (48.1) | 72 (36.5) | **0.018** |
| High | 10 (5.7) | 8 (7.8) | 0.5021 | 11 (5.3) | 2 (1.0) | **0.013** |
| Unknown | 12 (6.9) | 8 (7.8) | 0.782 | 14 (6.9) | 8 (4.1) | 0.217 |
| Smoking, n/N’ (%) |  |  |  |  |  |  |
| Active smoker | 50 (28.7) | 11 (10.8) | **<.001** | 37 (17.8) | 25 (12.7) | 0.154 |
| Ex smoker | 48 (27.6) | 29 (28.4) | 0.886 | 113 (54.3) | 60 (30.5) | **<.001** |
| Nonsmoker | 63 (36.2) | 54 (52.9) | **<.001** | 45 (21.6) | 105 (53.3) | **<.001** |
| Unknown | 12 (7.5) | 8 (7.8) | 0.928 | 13 (6.3) | 7 (3.6) | 0.211 |
| Alcohol, n/N’ (%) |  |  |  |  |  |  |
| Heavy drinker | 11 (6.3) | 0 (0.0) | **0.004** | 16 (7.7) | 3 (1.5) | **<.01** |
| Drinker, but not risky | 70 (40.2) | 26 (25.5) | **0.012** | 114 (54.8) | 60 (30.5) | **<.001** |
| Teetotal | 79 (45.4) | 68 (66.7) | **<.001** | 60 (28.8) | 122 (61.9) | **<.001** |
| Unknown | 14 (8.0) | 8 (7.8) | 0.953 | 18 (8.7) | 12 (6.1) | 0.318 |
| Alcohol units per week, median (interquartile range) | 0.4 (0.0-5.8) | 0.0 (0.0-75.0) | **<.001** | 2.0 (0.0-10.1) | 0.0 (0.0-0.7) | **<.001** |
| MEDAS Score *◊*, n/N’ (%) |  |  |  |  |  |  |
| Low adherence, 0-5 | 19/159 (11.9) | 3/93 (3.2) | **0.012** | 12/193 (6.2) | 5/186 (2.7) | 0.099 |
| Medium adherence, 6-10 | 109/159 (68.6) | 64/93 (68.8) | 0.974 | 131/193 (67.9) | 136/186 (73.1) | 0.267 |
| High adherence ≥11 | 31/159 (19.5) | 26/93 (28.0) | 0.126 | 50/193 (25.9) | 45/186 (24.2) | 0.703 |
| 14-MEDAS Questionnaire, n/N’ (%) |  |  |  |  |  |  |
| Olive oil as main source of fat | 147/161 (91.3) | 86/93 (92.5) | 0.745 | 190/194 (97.9) | 184/189 (97.4) | 0.706 |
| Consumption of more than 3 tablespoons of olive oil per day | 80/161 (49.7) | 42/93 (45.2) | 0.487 | 94/194 (48.5) | 96/188 (51.1) | 0.610 |
| Consumption of vegetables per day,  more than 1 portion | 75/161 (46.6) | 60/93 (64.5) | **<.01** | 97/194 (50.0) | 115/188 (61.2) | **0.028** |
| Consumption of fruits per day,  more than 2 | 75/161 (46.6) | 62/93 (66.7) | **<.01** | 116/193 (60.1) | 139/188 (73.9) | **<.01** |
| Consumption of red meat per day,  less than 1 | 102/160 (63.8) | 72/93 (77.4) | **0.024** | 147/194 (75.8) | 139/187 (74.3) | 0.745 |
| Consumption butter per day,  less than 1 | 115/160 (71.9) | 70/93 (75.3) | 0.557 | 139/194 (71.6) | 141/188 (75.0) | 0.459 |
| Consumption sugary drinks per day,  less than 1 | 105/161 (65.2) | 68/93 (73.1) | 0.193 | 132/194 (68.0) | 132/188 (70.2) | 0.646 |
| Consumption wine per week,  more than 2 glasses | 22/160 (13.8) | 3/93 (3.2) | **<.01** | 61/194 (12.4) | 12/188 (6.4) | **<.001** |
| Consumption legumes per week,  more than 2 portions | 95/160 (59.4) | 50/93 (53.8) | 0.384 | 119/194 (61.3) | 105/188 (55.9) | 0.276 |
| Consumption fish per week,  more than 2 | 92/160 (57.5) | 51/93 (54.8) | 0.681 | 115/194 (59.3) | 123/187 (65.8) | 0.190 |
| Consumption sweets per week,  less than 3 | 110/159 (69.2) | 73/93  (78.5) | 0.110 | 129/194 (66.5) | 137/188 (72.9) | 0.175 |
| Consumption nuts per week, at least 1 | 98/160 (61.3) | 53/93 (57.0) | 0.505 | 109/194 (56.2) | 87/188 (46.3) | 0.053 |
| Consumption preferably white meat per week | 123/160 (76.9) | 86/93  (92.5) | **<.01** | 142/194 (73.2) | 165/188 (87.8) | **<.01** |
| Consumption sautéed vegetables per week, more than once | 101/160 (63.1) | 56/93  (60.2) | 0.646 | 123/194 (63.4) | 123/188 (65.4) | 0.680 |

*Comparisons between groups were performed using the χ2 test for categorical variables and the t test for continuous variables. The Z test was used for comparisons between subcategories. Cardiovascular disease considers coronary heart disease, peripheral artery disease and cerebrovascular disease. n/N’: number of cases divided by total number of participants with available information. Bold values denote statistical significance.* ** The measure of variables that do not conform to a normal distribution is given in median (interquartile range) and comparison were performed using Mann-Whitney U test. ◊ MEDAS Score categories: Low adherence=0-5 points; Medium adherence=6-10; High adherence=11-15.*
